# Single-cell proteome profiling reveals distinct immunological patterns in the lungs of patients with severe acute respiratory failure

**DOI:** 10.64898/2026.01.06.26343420

**Authors:** Shiqi Zhang, Sebastiaan Joosten, Leonoor S Boers, Hélène B van den Heuvel, Tamara Dekker, Reece Davison, Juan J. Garcia Vallejo, Tom van der Poll, JanWillem Duitman, Lieuwe D J Bos, the BALI consortium

## Abstract

In this study, we provide a comprehensive characterization of the alveolar immune landscape in patients suffering from severe acute respiratory failure, predominantly caused by pneumonia or acute respiratory distress syndrome, conditions defined by intense pulmonary inflammation and immune dysregulation. Despite diverse underlying causes, the overall composition of alveolar immune cells was largely consistent, with neutrophils and macrophages comprising the majority of cells. However, the maturation and activation states of immune cell subsets varied significantly, not only between patients with and without pneumonia, but also among pneumonia cases stratified by pathogen type. We also observed dynamic shifts in immune cell subsets over the disease course and found that an increased proportion of CD123^bright^ immature neutrophils and a reduction in alveolar resident macrophages were associated with increased 28-day mortality. Integration with alveolar cytokine profiles revealed strong correlations between immune cell populations and the local cytokine milieu. These findings highlight the importance of assessing immune cell function, not merely abundance, through broad and longitudinal investigation to better understand the pathophysiology of acute respiratory failure and to guide precision immunomodulatory therapy.

## Introduction

Patients suffering from severe acute respiratory failure (SARF) often experience prolonged mechanical ventilation and extended stay in the intensive care unit (ICU), with a substantially increased risk of short-term mortality.(1–4) Pneumonia and acute respiratory distress syndrome (ARDS) are the most prevalent among the various causes of SARF.(3) Notably, pneumonia not only contributes directly to SARF but also stands as a primary risk factor for the development of ARDS.(5) These interrelated pathological states share core mechanisms marked by alveolar inflammation and dysregulated pulmonary immune responses, which culminate in tissue damage and the development of respiratory failure.(1, 6) Initial treatment focuses on addressing the underlying cause and providing supportive care.(5) However, by the time invasive mechanical ventilation is initiated, the causative pathogen is often already controlled or even eliminated, while ongoing alveolar hyperinflammation continues to cause lung injury.(6) At this stage, the therapeutic priority shifts toward resolving inflammation to promote tissue repair and support recovery, emphasizing the need to understand dynamic host immunology to guide targeted interventions.(5)

Because obtaining broncho-alveolar specimens is technically challenging, current insights into immune dysregulation in these patients are derived predominantly from peripheral blood, which has been shown to not accurately reflect the localized biological process occurring within the lungs because immune responses in pneumonia and ARDS are compartmentalized, with significant discrepancies between the alveolar milieu and systemic circulation.(7–9) Importantly, the pulmonary immune profile correlates more closely with disease severity and clinical outcomes than systemic measurements.(10, 11) Therefore, direct sampling of the alveolar compartment is essential to elucidate the immune mechanisms driving pulmonary pathophysiology in conditions such as ARDS and pneumonia.

Alveolar immune cells are pivotal regulators of immunoinflammatory responses within the lungs and play a central role in the progression of pneumonia and ARDS.(12–14) During this process, the alveolar compartment undergoes profound immunological remodeling, characterized by a marked influx of immune cells in response to pathogen invasion and increased alveolar-capillary permeability.(12) Unraveling the compositional and functional heterogeneity of these immune populations across distinct clinical trajectories is key to developing precise prognostic tools and interventions.

Previous studies on localized host responses have either relied on endotracheal aspirates, which reflect airway inflammation and offer limited cellular resolution for alveolar inflammation (15), or on bronchoalveolar lavage (BAL) performed in small cohorts analyzed through transcriptomic profiling, which fails to capture proteomic characteristics.(16) The COVID-19 pandemic, however, provided an unprecedented opportunity to study alveolar immune responses at high resolution, as large numbers of patients underwent BAL sampling and single-cell analyses during the peak of the crisis.(10, 11, 17–19) While these studies advance our understanding of immune cell dynamics in viral pneumonia, they are largely limited to SARS-CoV-2 cohorts and do not capture the broader landscape of alveolar immunity across different pneumonia etiologies. To date, comprehensive investigations of local immune cells at the protein level across a diverse SARF patient cohort remain lacking.

In this study, we aimed to characterize the composition and functional states of alveolar immune cells at the single-cell proteomic level, to describe differences across etiologies, and to quantify associations with clinical outcomes. We hypothesize that distinct immune cell subsets emerge depending on the presence or absence of pneumonia and the type of causative pathogen, and that patients with favorable clinical outcomes show a change in alveolar immune populations consistent with immune recovery. We also examined the connection between alveolar immune cells and the surrounding cytokine environment to understand how the inflammatory context might influence or mirror the functions and activation of immune cells.

## Results

### Patients

We included data from a total of 128 BAL fluid samples from 91 mechanically ventilated patients. Most patients were diagnosed with ARDS (81; 89%) and/or pneumonia (78; 86%). The most common cause of pneumonia was bacterial (bacterial pneumonia; 21/78; 27%), followed by viral pneumonia (14/78; 18%) and a combination thereof (bacterial+viral pneumonia; 10/78; 13%). The other 33 (42%) patients had no obvious pathogen identified (culture-negative pneumonia). Samples were collected at different time points and were well distributed across disease stages: 42 in the early stage (0–4 days post-intubation; 33%), 44 in the middle stage (5–10 days post-intubation; 35%), and 41 in the late stage (>10 days post-intubation; 31%). Overall, 67 (73.6%) patients survived 28 days post-intubation. (Figure S1, Table 1). All hereafter described analyses are focused on the presence of pneumonia and the causative pathogen (Table S1), mortality at day 28 (Table S2), taking the time point of sample collection into account (Table S3, Figure S2).

**Table 1.**
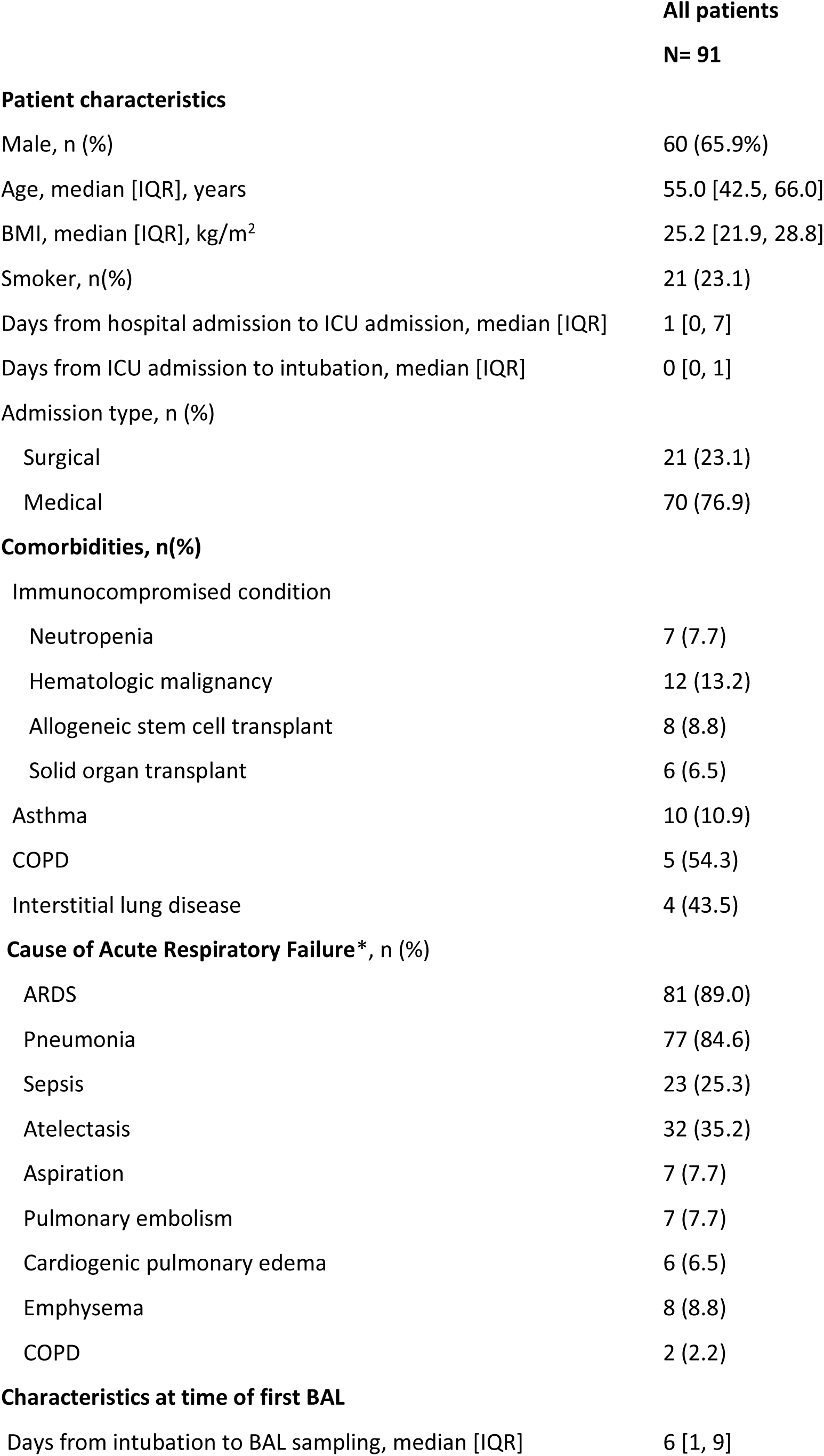
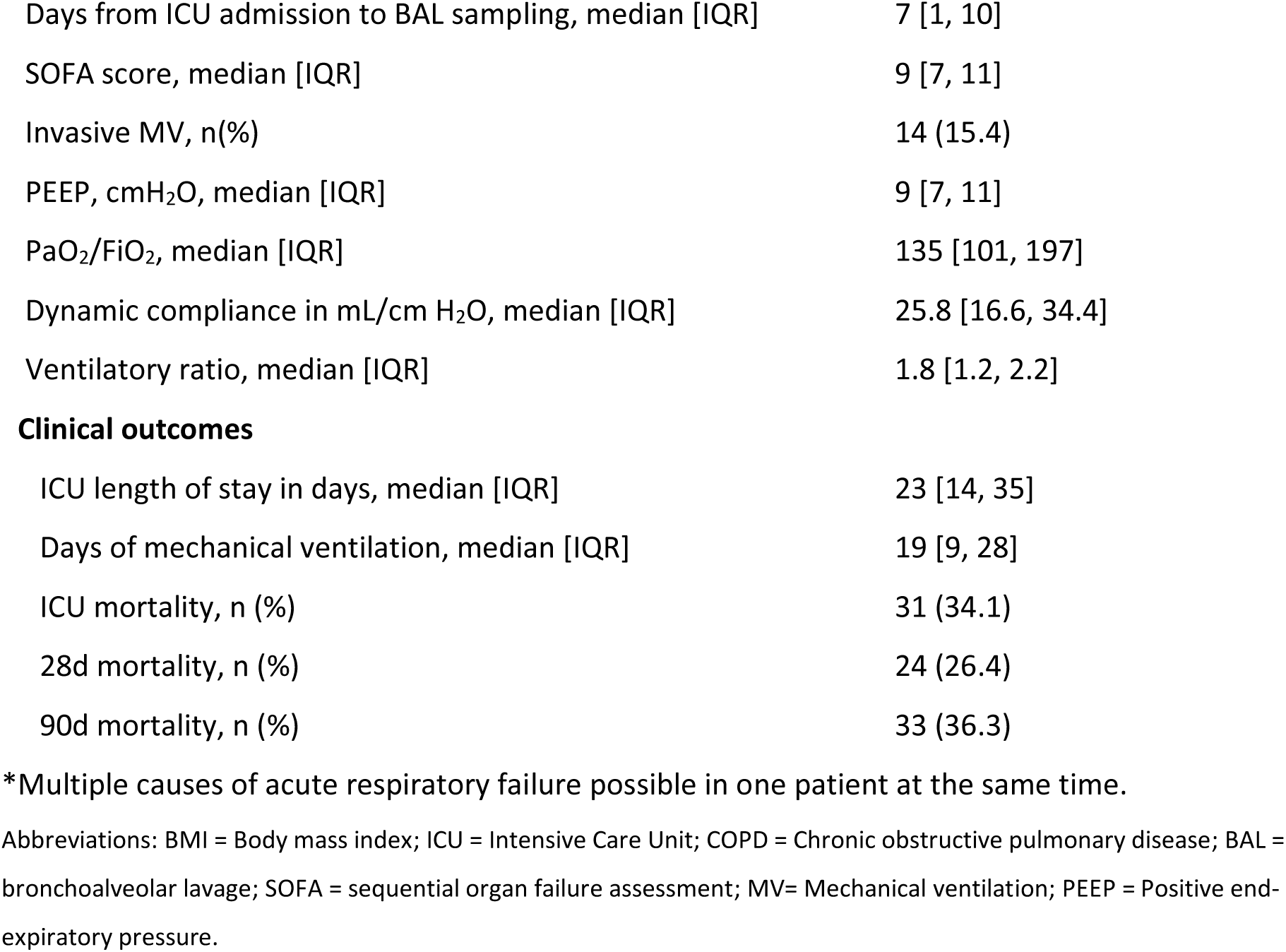
Demographics and clinical characteristics of the patients included in this study.

|  | **All patients**<br>**N= 91** |
| --- | --- |
| **Patient characteristics** |  |
| Male, n (%) | 60 (65.9%) |
| Age, median [IQR], years | 55.0 [42.5, 66.0] |
| BMI, median [IQR], kg/m<sup>2</sup> | 25.2 [21.9, 28.8] |
| Smoker, n(%) | 21 (23.1) |
| Days from hospital admission to ICU admission, median [IQR] | 1 [0, 7] |
| Days from ICU admission to intubation, median [IQR] | 0 [0, 1] |
| Admission type, n (%) |  |
| Surgical | 21 (23.1) |
| Medical | 70 (76.9) |
| **Comorbidities, n(%)** |  |
| Immunocompromised condition |  |
| Neutropenia | 7 (7.7) |
| Hematologic malignancy | 12 (13.2) |
| Allogeneic stem cell transplant | 8 (8.8) |
| Solid organ transplant | 6 (6.5) |
| Asthma | 10 (10.9) |
| COPD | 5 (5.4) |
| Interstitial lung disease | 4 (4.3) |
| **Cause of Acute Respiratory Failure\*, n (%)** |  |
| ARDS | 81 (89.0) |
| Pneumonia | 77 (84.6) |
| Sepsis | 23 (25.3) |
| Atelectasis | 32 (35.2) |
| Aspiration | 7 (7.7) |
| Pulmonary embolism | 7 (7.7) |
| Cardiogenic pulmonary edema | 6 (6.5) |
| Emphysema | 8 (8.8) |
| COPD | 2 (2.2) |
| **Characteristics at time of first BAL** |  |
| Days from intubation to BAL sampling, median [IQR] | 6 [1, 9] |
| Days from ICU admission to BAL sampling, median [IQR] | 7 [1, 10] |
| SOFA score, median [IQR] | 9 [7, 11] |
| Invasive MV, n(%) | 14 (15.4) |
| PEEP, cmH<sub>2</sub>O, median [IQR] | 9 [7, 11] |
| PaO<sub>2</sub>/FiO<sub>2</sub>, median [IQR] | 135 [101, 197] |
| Dynamic compliance in mL/cm H<sub>2</sub>O, median [IQR] | 25.8 [16.6, 34.4] |
| Ventilatory ratio, median [IQR] | 1.8 [1.2, 2.2] |
| **Clinical outcomes** |  |
| ICU length of stay in days, median [IQR] | 23 [14, 35] |
| Days of mechanical ventilation, median [IQR] | 19 [9, 28] |
| ICU mortality, n (%) | 31 (34.1) |
| 28d mortality, n (%) | 24 (26.4) |
| 90d mortality, n (%) | 33 (36.3) |

\*Multiple causes of acute respiratory failure possible in one patient at the same time.
Abbreviations: BMI = Body mass index; ICU = Intensive Care Unit; COPD = Chronic obstructive pulmonary disease; BAL = bronchoalveolar lavage; SOFA = sequential organ failure assessment; MV= Mechanical ventilation; PEEP = Positive end-expiratory pressure.

### Overview of the BAL fluid immune cell profile in SARF patients

BAL samples were collected from ICU patients with SARF and processed to obtain both cellular and supernatant fractions. A 50 marker CyTOF panel was applied to quantify major leukocyte lineages, cellular maturation, and activation states (Figure 1A & Table S4). Given the high proportion of debris inherent to BAL samples, an optimized clean up procedure was performed prior to downstream analysis (Figure S3). Cells were first partitioned into neutrophil, macrophage/monocyte, and lymphocyte compartments using FlowSOM, followed by higher resolution PhenoGraph clustering within each lineage.(20) Cluster identity was defined using marker enrichment modeling in conjunction with reference-based manual annotation. Dimensionality reduction with tSNE and PacMAP confirmed consistent topological organization across samples, and marker expression profiles are detailed in Supplementary Figures S4–S7.

**Figure 1.**
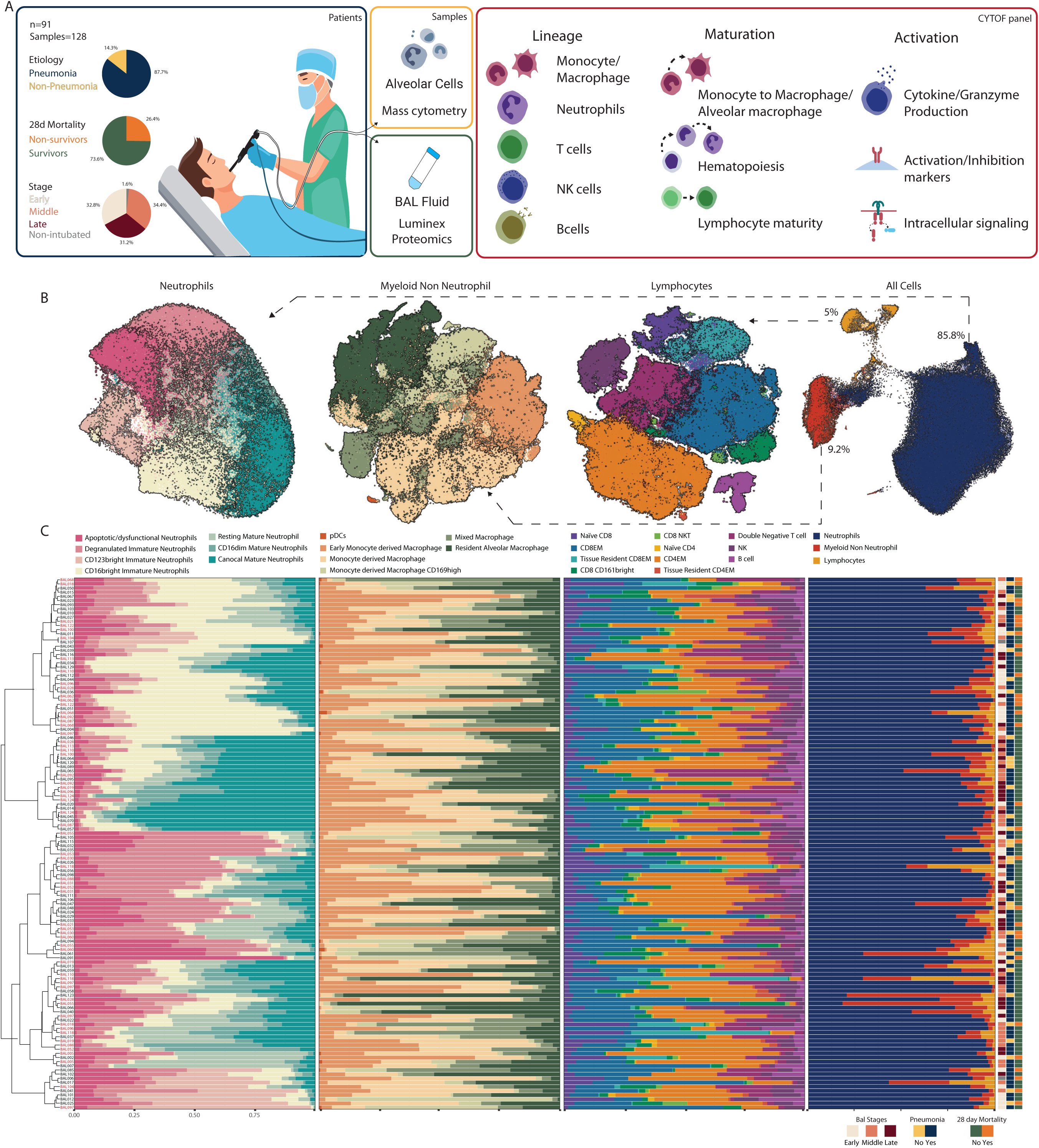
Immune cell composition in BALF samples from all patients. Panel A: Overview of the study design, patient cohort characteristics, sample processing and the CyTOF panel. Panel B: Dimensionality reduction using PaCMAP reveals three major cell types: neutrophils, myeloid non-neutrophils and lymphoid cells. Panel C: Further clustering within each cell types is shown on the left. Frequency distribution of immune cell subsets across individual BAL samples, hierarchically clustered using neutrophil cluster abundance. Samples are stratified by key clinical parameters, including disease stage (early 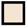, middle 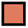, late 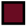), pneumonia diagnosis (yes 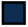 /no 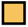), and mortality at 28-day post-intubation (no 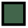/ yes 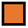).

Across the cohort, the BAL cellular landscape exhibited a strikingly conserved composition. Neutrophils dominated the compartment, comprising a mean of 85.8% of CD45+ leukocytes, followed by macrophages (9.2%) and lymphocytes (5.0%) with limited variation across etiologies or sampling time (Figure 1B-C; right column). These highly consistent proportions indicate that clinically meaningful differences are more likely to arise from variation in cellular phenotype rather than overall abundance.

Despite their uniform dominance, neutrophils showed substantial phenotypic diversity. Maturation status was characterized by graded expression of CD10 and CD16, revealing a broad continuum rather than discrete developmental subsets. A distinct subset of immature neutrophils (Figure 1B-C; left column) displayed elevated CD123 (IL-3Rα), suggesting divergent programming. Almost all neutrophils bore features of tissue activation, including downregulation of CD62L following alveolar recruitment. These observations underscore the functional diversification of the neutrophil response in respiratory failure. Samples were clustered based on the relative abundance of neutrophil subsets (Figure 1C; left column).

Macrophages represented the second largest immune cell group in BAL. In health, the alveoli are predominantly populated by resident macrophages, while in infection or ARDS, monocyte derived macrophages infiltrate the lung to support pathogen control and tissue remodeling.(21) Here, high dimensional projections showed no sharp separation between resident and recruited subsets but rather a gradual phenotypic continuum, consistent with progressive acquisition of resident and inflammatory like traits by incoming monocytes.

Markers of resident identity (CD206, CD71, CD169) and monocyte origin (CD14) were used to infer ontogeny, complemented by CD163, CD86, and HLA-DR to capture functional states (Figure 1B).(21–24) Notably, markers commonly used to distinguish anti-inflammatory versus pro-inflammatory macrophages such as CD206, CD163 and CD86 were frequently co expressed, indicating that simple classifications into pro- or anti-inflammatory types do not adequately capture the diversity of alveolar myeloid cells (Figure S4A).(25)

Lymphocytes formed a smaller but highly heterogeneous cell population, comprising 11 distinct clusters. Effector memory CD4+ T cells and cytotoxic CD8+ T cells were prominent across disease etiologies, indicative of a robust antigen experienced response within the alveolar space in most patients.

Additionally, we identified that lymphocyte subsets were rarely observed in peripheral blood. Double negative T cells (CD4− CD8−) expressed high levels of granzyme B and perforin, resembling cytotoxic T cells lacking CD8 expression.(26, 27) We further identified a CD161^bright^ cytotoxic population with innate like features. The abundance of these specialized phenotypes varied markedly between patients.(28, 29)

### Immune cell composition and activation are influenced by pneumonia and the causative pathogen

The host immune response is a critical determinant of pneumonia progression and resolution, with immune dynamics playing a key role in shaping disease trajectory.(30) We first compared the composition of alveolar immune cells between patients with and without pneumonia. Using Hedge’s *g* to quantify effect sizes,(31) we observed no significant differences in either major immune cell types or their subsets between patients with and without pneumonia (Figure S8).

Given the heterogeneity of pneumonia and the critical role of the causative pathogen in shaping alveolar immune responses and influencing clinical outcomes (32), we assessed differences in the composition of alveolar immune cells stratified by pathogen type (Figure 2A: bacterial, viral or combined pneumonia vs no causal pathogen identified).(33) We performed principal component analysis (PCA) on activation markers relevant to specific cell types (Figure 2B). Markers that were highly correlated, as indicated in the PCA and represented with similar colors, were used to construct a score indicative of that specific activation pattern as seen in Figure 2A.(34) Together, these analyses cover the differences in composition and activation between causative pathogens for pneumonia.

**Figure 2.**
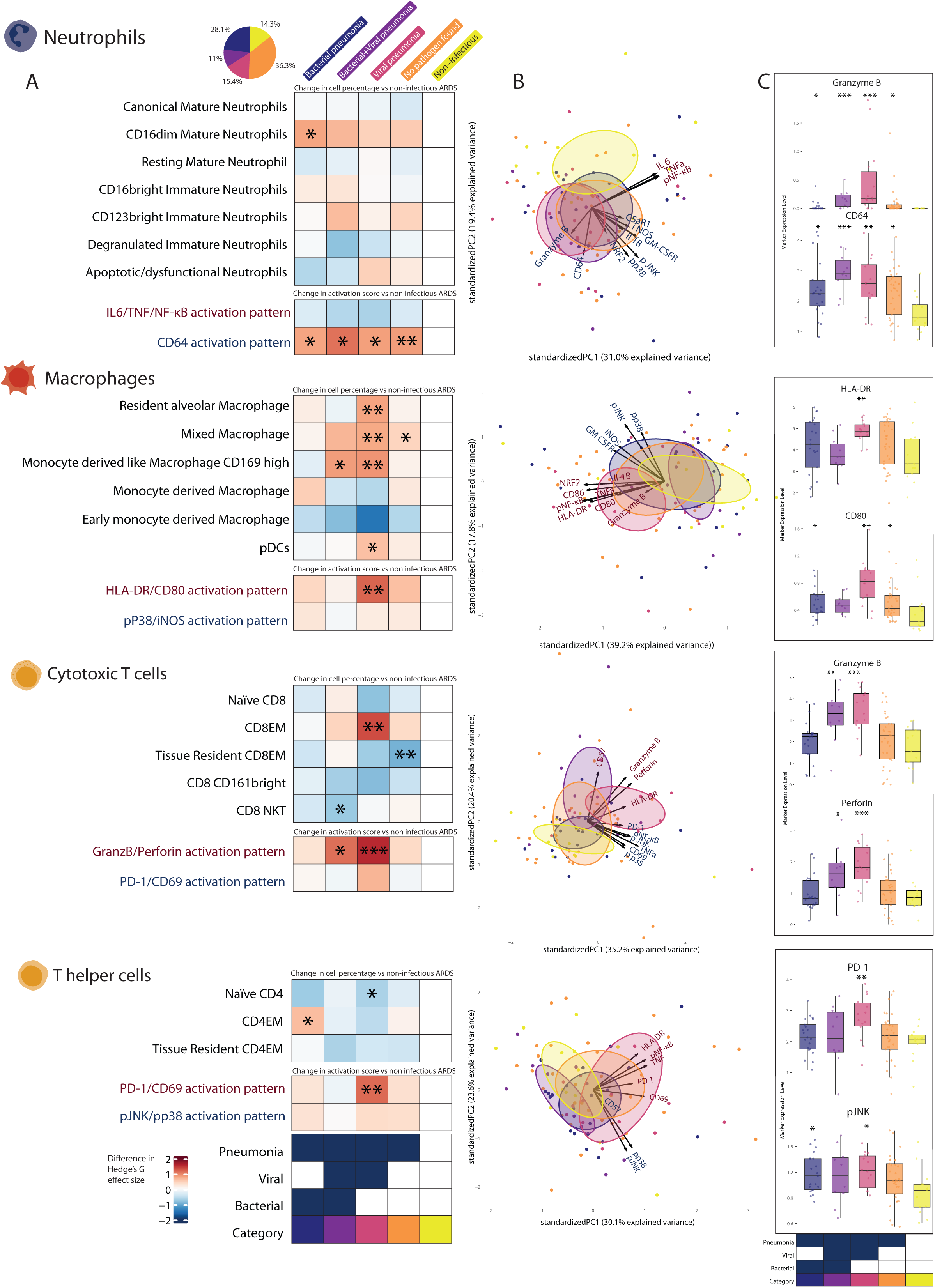
Alveolar immune cell subsets and distinct activation patterns across pneumonia etiologies. Panel A: Heatmap showing changes in cell subset abundance and activation scores across bacterial, viral, combined bacterial and viral pneumonia as well as culture negative pneumonia and non-pneumonia patients. Each box indicates the effect size relative to the non-pneumonia group. Statistically significant changes are denoted by asterisks (*p < 0.05, **p < 0.01, ***p < 0.001). Panel B: Principal component analysis (PCA) of functional marker expression that differentiates pneumonia groups. Vectors represent the contribution of individual markers to group separation. Markers that were highly correlated are represented with similar colors. Panel C: Comparisons among the expression levels of key functional markers across different pneumonia groups. Color boxes represent pneumonia subgroups: blue 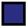 for bacterial pneumonia, purple 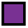 for bacterial+viral pneumonia, pink 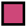 for viral pneumonia, orange 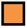 for culture-negative pneumonia, yellow 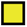 for non-pneumonia.

Neutrophils have been reported as the most prevalent immune cell in the lungs of patients with bacterial and viral pneumonia. In our analysis, we identified an increase in CD16^dim^ mature neutrophils in patients with bacterial pneumonia, compared to patients without pneumonia. Loss of CD16 in mature neutrophils indicates senescence programming and may reflect decreased phagocytotic properties.(29, 35) Alternatively, CD16 can be lost upon activation of mature neutrophils. Our current analysis cannot differentiate between the two, but there was no association with age (P=0.48), making the latter more likely. Neutrophil activation patterns, characterized by cellular CD64, NRF2, p-p38, p-JNK, and IL1B content, was increased in all patients with pneumonia. CD64 is also known as FcγRI, an interferon-inducible, high-affinity IgG receptor and is a well-established marker for neutrophil activation in bacterial infections and can serve as a prognostic marker in septic shock.(36–39) The type of pathogen did not appear to substantially affect the immune response, as these effects were largely consistent between causative pathogens. This aligns with the classical view of neutrophils as non-specific effectors of infection with a broad, non-targeted response. Previous studies have demonstrated neutrophil infiltration in both bacterial and COVID-19 pneumonia, without in-depth analysis of their activation patterns.(40–42) Our findings expand the current understanding, not only are neutrophils the most common cell in all types of pneumonia, but neutrophil activation signatures are also similar across broad categories of causative pathogens.

Alveolar macrophages (AMs) are largely replaced by monocyte-derived macrophages (MDMs) as the dominant macrophage population in the lungs of patients with severe respiratory failure caused by viral infections, mostly due to SARS-CoV2 but also in a preclinical Influenza model.(23, 41, 43, 44) We confirmed an increased abundance of MDMs in patients with viral pneumonia, but simultaneously observed a higher abundance of mixed macrophages, with lineage markers of monocyte as well as AMs origin making their origin uncertain, and of alveolar macrophages in the same patients. While a collapse of the resident AM network has previously been described in COVID-19 related respiratory failure (45) we did not observe a significant decline in this population among the viral pneumonia patients in our study. Our results indicate that there is a highly heterogeneous monocyte/macrophage response to viral pneumonia, balancing between homeostatic AMs and MDMs with distinct pathogenic states and that the collapse described in COVID-19 occurs in some patients, but not all. (23)

CD8 effector memory (CD8EM) cells were also more abundant in the lungs of patients with viral pneumonia, compared to the other causative pathogens. We postulate that this long-living cell population with antigen-specific function maintains local host response against the viral pathogen. Its abundance may reflect the timing of sampling of the included patients, most patients with viral pneumonia admitted to the ICU were likely infected more than a week before, when the adaptive host response is well-established. In line with that, we identified increased activation of both CD4+ helper T cells and CD8+ cytotoxic T cells in viral pneumonia patients. Activation of CD8+ cytotoxic T cells in viral pneumonia was defined by increased protein content of Granzyme B, perforin, HLA-DR and CD57. CD4+ helper T cells had increased programmed cell death protein 1 (PD-1), HLA-DR, CD69, tumor necrosis factor (TNF) and Nuclear factor-kB (NF-kB) content. Granzyme B-associated activation patterns, reflecting antiviral effector programming (46), were elevated across all adaptive immune lineages in viral pneumonia patients. This broad cytotoxic activation partially overlaps with previous findings in SARS-CoV-2 infection, where such responses were largely confined to cytotoxic lymphocytes and were associated with severe comorbidity.(47, 48) The response also aligns with the increased HLA-DR expression found on CD4+ and CD8+ lymphocytes and heightened CD69 abundance on CD4+ helper T cells. Simultaneously, we found evidence for immune balance PD-1 activation pattern, indicating sustained local activation and a potential progression towards functional exhaustion, which was previously observed in chronic inflammatory disease.(49) Together, these results underscore pathogen-specific immune remodeling in pneumonia patients, driven by distinct cellular response and signaling pathway dynamics.

### Temporal dynamics of alveolar immune cell subsets are associated with patient clinical outcomes

To understand how alveolar immunity evolves during SARF, we conducted joint longitudinal and survival modelling, combining linear mixed effects and Cox proportional hazard modelling. Joint models have specific advantages necessary for this dataset; they can handle irregular sampling times commonly encountered in studies relying on alveolar sampling, estimate the longitudinal effects (e.g. the change in a subset of cells over time) while controlling for informative censoring due to death and quantify the risk of death related to changes in cell composition. These models provide an estimate of the change in 10% abundance of a cell population, or activation score per additional day on mechanical ventilation as well as a hazard ratio for death for a similar range.

Using this approach, we identified a significant decline in two immature neutrophil subsets: CD16^bright^ and CD123^bright^ (Figure 3; neutrophil panel – left side: Longitudinal model), while degranulated immature neutrophils were stable over time. Emergency myelopoiesis is well-described in sepsis and pneumonia and typically observed in blood.(50) These cells are subsequently recruited to the lungs, where they accumulate and colocalize with areas of diffuse alveolar damage.(51, 52) Even though immature subsets decrease with time, their abundance was still non-neglectable, even more than a week after the initiation of mechanical ventilation (approximately 50% of neutrophils). This contrasts with the presence of immature neutrophils in blood, which typically disappear within days.(53) The persistence of alveolar neutrophils over this time span can only be explained by continued recruitment of neutrophils as their lifespan is several days according to the most liberal estimations.(54) Recruitment of CD16^dim^ mature neutrophils increased their abundance over time and is associated with a decrease in mortality (Figure 3; neutrophil panel – right side: Survival model). Even though CD16 can also be considered a maturity marker, we classified these cells as mature neutrophils based on their CD10^bright^ status. We will give additional evidence and details that supports this choice in the section on *trajectory and morphology of alveolar neutrophils*.

**Figure 3.**
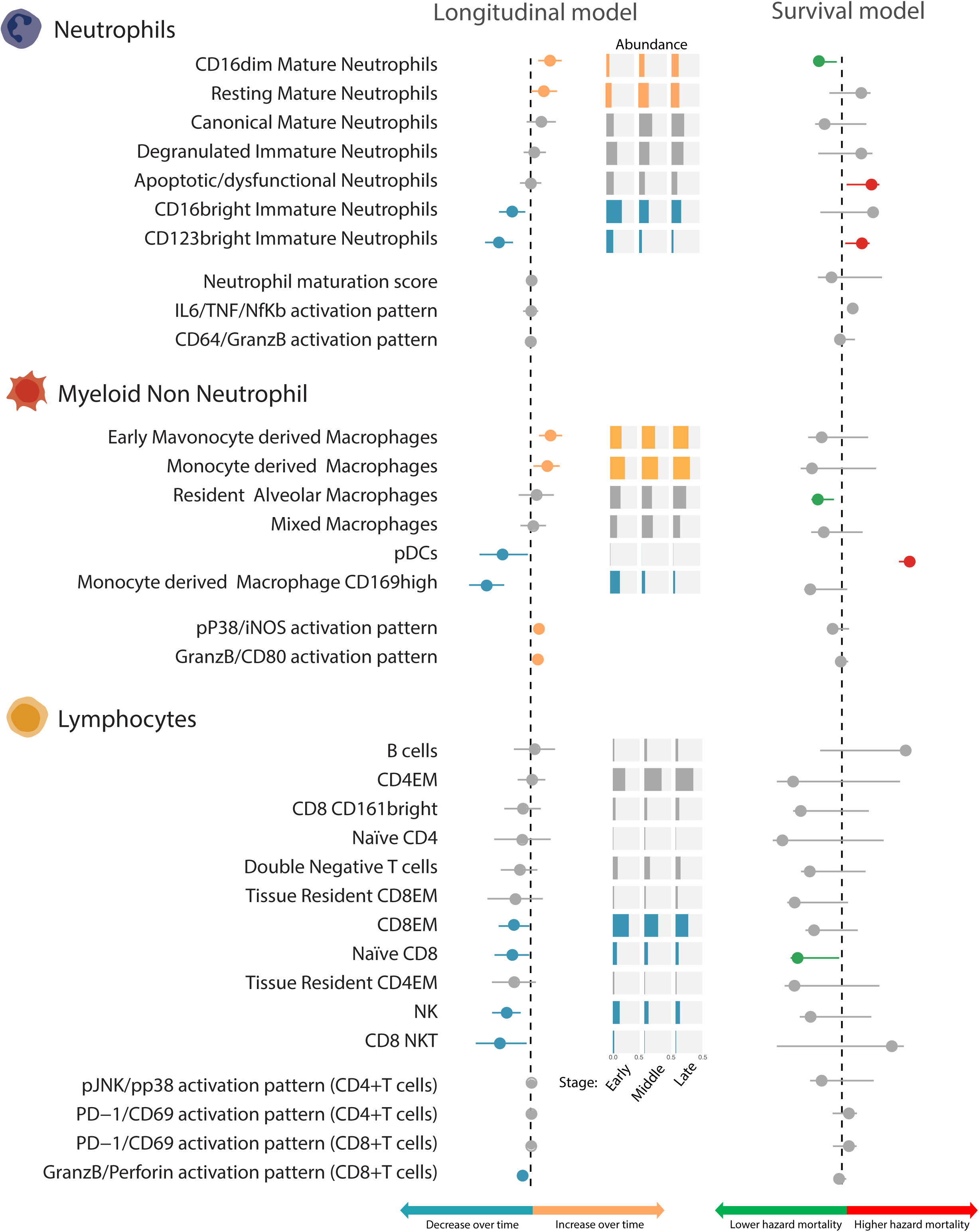
Dynamic changes in immune cell subsets and activation scores and their associations with 28-day mortality. Left: Longitudinal model shows the temporal trends in cell subset abundance or activation score over time. A **yellow line** indicates a significant increase over time, while **blue lines** indicate a decrease over time. Middle: Stacked bar plots display the relative proportions of each subset within its parent lineage (neutrophils, myeloid non-neutrophil cells or lymphocytes) at different disease stages (early / middle / late). Colors are similar to the results from the longitudinal model. Right: The survival model evaluates the relationship between each subset and 28-day mortality. A **red line** indicates that increased abundance is associated with a higher risk of death, **green lines** indicate an association with lower mortality risk. **Gray lines** represent non-significant findings.

Immature neutrophils have previously been linked to systemic hyperinflammation and myeloid dysfunction in sepsis and ARDS. (55–57) Here we identified a strong association between a higher percentage of CD123^bright^ immature neutrophils and mortality. CD123, also known as IL-3 receptor, is expressed on immature neutrophils in the bone marrow and allows for rapid recruitment of these cells through upon local release of IL-3 in the lungs, a pathway independent of Granulocyte-Colony-Stimulating Factor (GCSF). These cells were previously identified in the blood of patients with sepsis and COVID19, and are associated severity of disease and hyperinflammation.(58, 59) We speculate that this particular subset of immature neutrophils is constantly released from the bone marrow and recruited into the alveolar compartment through IL-3 release during uncontrolled local hyperinflammation.(58, 60)

We observed a shift from CD169^bright^ MDMs to monocyte and early MDMs with a longer duration of mechanical ventilation (Figure 3; macrophages panel – left side). The CD169^bright^ cells share surface proteins with the resident AMs, while the (early) MDMs are more similar to classical monocytes. This suggests sustained monocyte recruitment without further differentiation into an AM phenotype. This phenomenon has been observed in both COVID-19 and non-COVID-19 ARDS,(22, 41, 61) indicating that it is not unique to SARS-CoV-2 infection and has previously been found to be associated with prolonged alveolar inflammation across ARDS etiologies. In line with this hypothesis, activation patterns related to p38/iNOS and to HLA-DR/CD80 increased over time, paralleling the changes in (early) MDMs. None of the MDM populations were, however, associated with mortality in our cohort. In contrast, a higher proportion of AMs was associated with a lower risk for death, possibly reflecting their regulatory role within the alveolar compartment (Figure 3; macrophages panel – right side) This interpretation is supported by the lower cytokine levels linked to this population (Supplementary Figure S9) and prior reports.(62) We also identified a decrease in plasmacytoid dendritic cells (pDCs) over time and a strong association of pDCs with higher mortality. This signal needs to be interpreted in the light of the very low abundance of these cells (median 0.1% of cells; Table S6) and uncertainty surrounding the estimation as a result of that. The cells are classified as pDCs rather than DCs because of their consistent CD123+ and CD11c- status (Figure S6).(63)

CD8EM cells were most abundant early in the disease course but decline significantly over time, mirroring the waning antiviral response of CD169^bright^ macrophages and a reduction in Granzyme B/perforin activation pattern. While the initial high abundance of these cells could reflect an early rush of infiltration into the lung, this recruitment tapered off and was not sustained. Consequently, our data suggests that although early sequestration may contribute to the onset of systemic lymphopenia often seen in patients with sepsis(64), the persistence of this condition is driven by lymphocyte exhaustion and apoptosis rather than ongoing accumulation in the alveolar space.

### Alveolar neutrophils maturity follows a trajectory and is reflected in cell morphology

Given the association of multiple immature neutrophils subsets with patient outcomes, we explored the maturation trajectory using pseudotime analysis by constructing a continuous trajectory of neutrophil differentiation with the Wanderlust algorithm (Figure 4A). This analysis incorporated a selection of canonical maturation markers (Table S5), allowing high-resolution ordering of individual neutrophils along a maturation continuum. The apoptotic or otherwise dysfunctional neutrophil cluster, characterized by aberrant expression profiles incompatible with normal maturation dynamics, was excluded from modeling.

**Figure 4.**
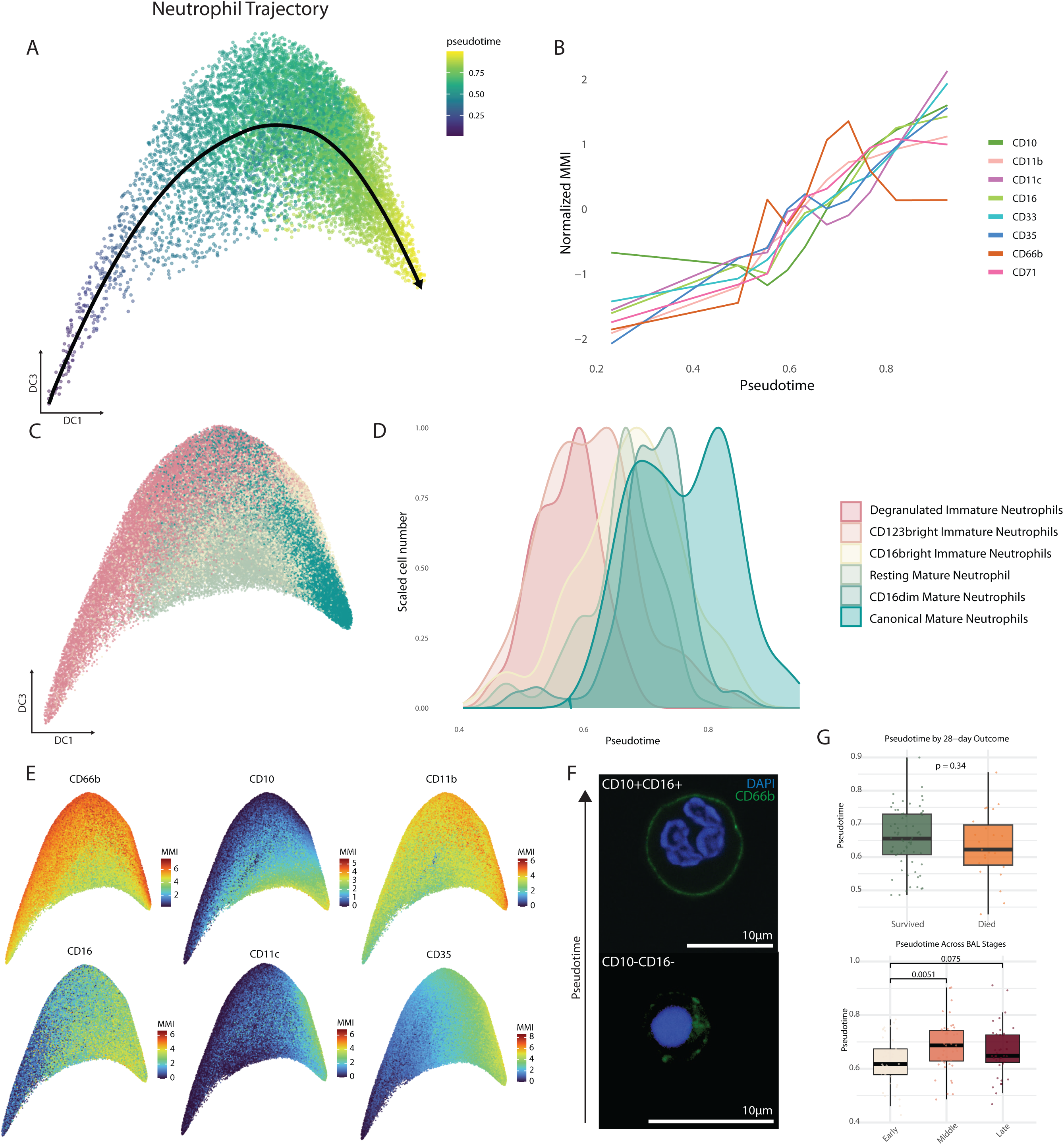
Neutrophil trajectory and maturation continuum based on pseudotime analysis. Panel A: Diffusion map showing the inferred neutrophil maturation trajectory, colored by pseudotime from immature (**purple**) to mature (**yellow**) states. Panel B: Expression dynamics of selected surface markers plotted along pseudotime. Panel C: Diffusion map colored by neutrophil subpopulation identity. Panel D: Density distribution of annotated neutrophil subsets along pseudotime. Panel E: Diffusion maps showing marker intensity if representative maturation and activation markers across pseudotime. Panel F: Representative immunofluorescence images of CD10^+^CD16^+^ and CD10^-^CD16^-^ immature neutrophils stained for **CD66b** and **DAPI**, illustrating morphological differences. Panel G: Boxplots showing pseudotime distribution in patients stratified by 28-day outcome.

The pseudotime trajectory revealed a coherent gradient of maturation, defined by increasing expression of CD10, CD16, CD11c, and CD35 as pseudotime advanced. CD66b expression remained relatively constant, as CD66b is an activation and not a maturation marker in sepsis (Figure 4B, E).(65, 66) Projection of the previously defined PhenoGraph clusters onto the pseudotime continuum confirmed strong agreement between pseudotime and phenotypically annotated subsets (Figure 4C, D).

To validate these findings, we performed fluorescence-activated cell sorting (FACS) on BAL samples from patients with either high levels of the canonical mature neutrophil subset or the degranulated immature subset. Neutrophils corresponding to the extremes of the pseudotime spectrum were isolated by sorting for high versus low CD10/CD16 expression. Cells within the CD10+CD16+ gate, representing the mature end of the trajectory, displayed the characteristic segmented morphology of terminally differentiated neutrophils. In contrast, CD10-CD16- cells were smaller with a predominantly mononuclear appearance, consistent with immature neutrophil morphology. No difference in overall CD66b expression was observed between both populations, confirming that activation and maturation represent distinct dimensions of neutrophil heterogeneity in the lung and validating our interpretation of neutrophil maturity and their presence within the lungs of these patients (Figure 4F, Figure S9A). Quantitative comparison of marker intensities between FACS and CyTOF data demonstrated strong concordance across platforms (Figure S11).

We next integrated neutrophil pseudotime into the longitudinal joint model to assess how alveolar neutrophil maturity evolves over the disease course and whether it relates to clinical trajectories (Figure 3; neutrophil maturation score). We further examined the median pseudotime of neutrophils in patients within the joint model and found no meaningful differences in patient outcomes, implying that specific neutrophil subsets are likely more relevant to clinical outcomes than neutrophil maturity alone. However, pseudotime values increased progressively with disease duration, indicating a temporal shift toward more mature neutrophil phenotypes in later disease stages. The lowest pseudotime values were consistently observed in early-stage samples, likely reflecting the surge of immature neutrophils entering the alveolar space during periods of intense emergency myelopoiesis early in disease progression. (16, 50)

### Local cytokine and chemokine signatures mirror alveolar immune cell profiles

The behavior of alveolar immune cells is shaped not only by pathogen-derived stimuli but also by the surrounding inflammatory milieu, which modulates their activation and function.(67, 68) We measured a broad range of protein biomarkers related to inflammation (pro and anti), immune cell activity, programmed cell death, coagulation and fibrinolysis, endothelial injury and epithelial injury to explore the relationship between local soluble protein responses and cellular composition and activation (Figure 5 and Table S7). Classical pro-inflammatory cytokines such as interleukin 1 beta (IL-1β), IL-33, and TNF were strongly correlated with neutrophil abundance and activation in the alveolar space, a hallmark of tissue-damaging inflammation in severe lung injury.(69, 70) Chemokines with neutrophil attractant properties, such as CCL3, CXCL2 and GCSF, were strongly correlated with the abundance of neutrophils, indicative of continuous recruitment. Neutrophil products such as matrix metalloproteinase-8 (MMP8), also known as neutrophil collagenase, were significantly correlated with neutrophil abundance, reflecting the degradation of extracellular matrix by extensive neutrophil driven inflammation in the lung. Neutrophil abundance was associated with a decrease in alveolar concentrations of surfactant protein D (SP-D), which is produced under homeostatic circumstances by alveolar type II cells. A decrease in SP-D is reflective of alveolar damage and results in the increase of alveolar surface tension and decreased pulmonary compliance, a hallmark of ARDS.

**Figure 5.**
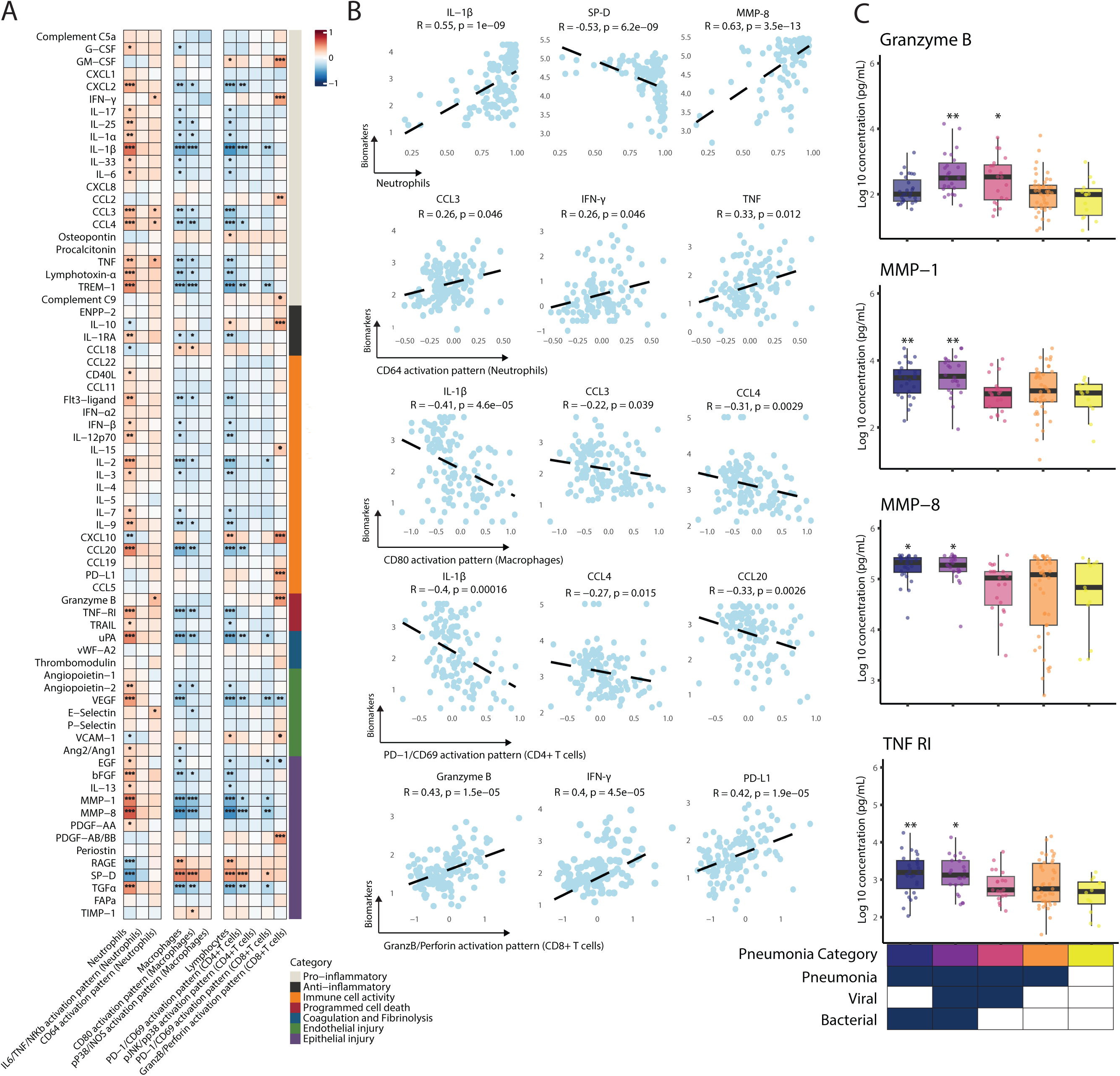
Alveolar cytokine associations with immune cell profiles and pneumonia etiologies. Panel A: Heatmap showing Spearman correlation coefficients between alveolar cytokine levels and immune cell abundances and activation scores in each immune lineage. Positive correlations are shown in **red** and negative correlations in **blue**. Cytokines are listed on the y-axis, and immune cell parameters are groups by lineage on the x axis. Panel B: Representative scatter plots showing significant correlation between selected cytokines and immune cell parameters, with Spearman correlation coefficients (R) and corresponding *p*-values indicated. Panel C: Boxplots showing the comparison of selected alveolar cytokine concentrations among patient categories. Color boxes represent pneumonia subgroups: blue 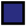 for bacterial pneumonia, purple 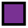 for bacterial+viral pneumonia, pink 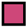 for viral pneumonia, orange 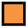 for culture-negative pneumonia, yellow 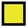 for non-pneumonia.

Macrophages and lymphocyte abundance showed inverse correlations to the neutrophils. They were correlated with lower concentrations of most of the pro-inflammatory cytokines and were correlated with higher alveolar SP-D concentrations. More activation of the Granzyme B/perforin signal on CD8+ T-cells was positively correlated with the soluble Granzyme B concentration, in line with the available literature indicating CD8+ T-cells as the primary producers of Granzyme B.(71) The activation was also correlated with increased concentrations of interferon (IFN)y. Both are important for viral control and are higher in viral pneumonia. IFNy can further regulate the cytotoxic behavior of CD8+ T-cells and modulate the immune response. Interestingly, activation of CD8+ T-cells was correlated with PD-L1 levels. PD-L1 serves to reduce the cytotoxic properties of these cells when bound to PD-1, suggesting an active negative feedback loop. Such a response can be physiological, limiting tissue injury, but could also be pathological resulting in evasion of the immune system by the pathogen through T-cell exhaustion. Surprisingly, the GM-CSF concentration was also strongly correlated with activation of CD8+ T-cells, yet a causal immunological relationship is unlikely due to their unrelated origin and mechanisms of action.

CD80 driven activation of macrophages should increase pro-inflammatory cytokine production (72, 73). Yet, we found an inverse correlation with IL-1β levels. PD-1 activation on CD4+ T cells was also associated with lower concentrations of several pro-inflammatory cytokines, such as IL-1b. However, canonical changes related to PD-1 activation, such as decreased IL-2 and IFNy production, were not replicated in the correlation analyses. Although CD69 can have pro and anti-inflammatory effects, co-expression of PD-1 and CD69 on CD4+ T-cells suggests an immunoregulatory role. The Granzyme B/Perforin activation pathway in CD8+ cytotoxic T cells was significantly correlated with cytokines involved in T cell activation, cytotoxicity, and exhaustion, including GM-CSF, Granzyme B, and PD-L1.(71)

We next compared the alveolar cytokine concentrations across patient groups (Figure 5C and Figure S12). Patients with bacterial or bacterial-viral co-infections showed elevated levels of key inflammatory mediators, including MMP-1, MMP-8, and Granzyme B.(74, 75) The increased levels of MMP-1 and MMP-8 are consistent with prior evidence implicating neutrophil-derived metalloproteinases in extracellular matrix degradation and alveolar-capillary barrier disruption in bacterial pneumonia and ARDS.(76, 77) In contrast, patients with viral pneumonia exhibited elevated Granzyme B levels, reflecting robust cytotoxic lymphocyte activity, consistent with previous reports on influenza and RSV infections.(78, 79) Taken together, these results demonstrate that local cytokine profiles closely mirror immune cell states, underscoring the importance of integrated cellular and cytokine analyses to fully characterize the immune response in pneumonia/ARDS.

## Discussion

Here, we provide a comprehensive immune cell atlas of the alveolar compartment of an etiology-stratified SARF cohort by applying high-dimensional mass cytometry with a 50-marker panel to longitudinal BAL samples. We show that, although overall immune cell composition remains relatively stable across patients, distinct etiologies, particularly pneumonia and pathogen type, reprogram immune cell functional states. These shifts manifest as dynamic changes in specific immune phenotypes over time, are associated with patient risk for death, and are mirrored by local cytokine patterns. Together, these findings reveal marked immunological heterogeneity in pneumonia and ARDS and identify potential immune targets tailored to defined patient subgroups.

A key contribution of our study is the detailed characterization of alveolar immunity across distinct etiologies in SARF patients, including various causes of pneumonia and ARDS. The striking stability in immune cell proportions suggests that clinically relevant differences arise instead from functional remodeling, which has received increasing attention in recent years, particularly during the COVID-19 pandemic.(10, 19, 80) Pathogen-specific activation of alveolar immune cells in pneumonia has been underexplored in earlier studies, many of which grouped pneumonia cases together without stratifying by etiology or lacked the analytical resolution to define functional states at the single-cell level.(7, 33) In contrast, our study offers a more nuanced perspective by directly comparing distinct pneumonia subtypes to non-pneumonia cases, enabling a pathogen-resolved characterization of the alveolar immune landscape, the substantial heterogeneity in the distribution of immune cell subsets across pathogen-defined subgroups suggests that the identity of the causative pathogen, rather than the mere presence of pneumonia, is the principal determinant shaping alveolar immune activity.

The longitudinal structure of our cohort enabled us to dissect how these phenotypes evolve during mechanical ventilation and how such trajectories are associated with survival. Several studies have demonstrated the recruitment of immature neutrophils into the lungs during the acute phase of severe COVID-19.(19, 81, 82) In our cohort, we observed a similar early enrichment of immature neutrophils and extended these findings by documenting a subsequent accumulation of mature subtypes at later disease stage. Notably, CD123^bright^ immature neutrophils were strongly associated with mortality, aligning with prior observations of IL-3-responsive immature granulopoiesis in systemic hyperinflammation.(58, 59) Crucially, the association between persistent immature neutrophils, specifically within the alveolar niche, and poorer prognosis is reported here for the first time in pneumonia/ARDS patients. These findings broaden our understanding of immature neutrophils, extending their relevance from the systemic circulation to localized immune responses within the lungs. Their association with poor outcomes suggests either a direct pathogenic role or a failure of maturation that compromises effective immune defense, both of which require further investigation. Integration with single-cell transcriptomics may clarify neutrophil maturation trajectories, similar to patterns recently characterized in mice.(83)

Macrophages play an orchestrating role in lung inflammation due to their remarkable plasticity and capacity to adapt to the local microenvironment following lung injury.(13, 84) A prior CyTOF analysis identified alveolar macrophage phenotypes associated with clinical outcomes in ARDS patients.(85) Building on this, our study demonstrates both substantial heterogeneity in macrophage activation states across pneumonia etiologies and a dynamic continuum from resident-like alveolar macrophage phenotypes towards canonical MDMs during SARF. This continuum indicates that macrophages transition not between discrete phenotypes but along a spectrum shaped by the inflammatory alveolar milieu.(86) Notably, the finding that resident-like macrophage phenotypes were associated with more favorable clinical trajectories raises the possibility that early depletion or reprogramming of this resident pool may diminish key homeostatic functions including efferocytosis, regulation of epithelial repair, and attenuation of excessive neutrophilic inflammation.(87, 88)

Interestingly, the cytokine microenvironment recapitulated, and likely helped drive, the immune gradients we observed at the cellular level. Neutrophil-recruiting cytokines such as CCL3 and CXCL2 rose in parallel with neutrophil accumulation, indicating ongoing waves of neutrophil recruitment into the alveoli, most likely driven by signals from activated monocytes or MDMs.(89–91) Because neutrophils are too short-lived to mature within the lung, these cytokine patterns support the interpretation that newly generated, bone marrow-derived neutrophils are continually attracted into the injured alveolar space. In contrast, cytokines associated with resident macrophage-mediated tissue-repair programs (e.g., IL-10, CCL18) correlate with lower neutrophil abundance, suggesting that homeostatic macrophage activity shifts the cytokine milieu toward reduced neutrophil recruitment and enhanced tissue protection.(92) Under high-inflammatory cytokine conditions, effector programs were upregulated in neutrophils and cytotoxic T cells, whereas activation of macrophages and T helper cells was suppressed.(70, 93) Together, these correlations underscore a bidirectional interplay in which immune cell programming both shapes and responds to the local mediator milieu. Such tightly linked cellular-cytokine networks may form feedback circuits that either promote effective pathogen clearance or, when dysregulated, drive excessive inflammation and subsequent tissue damage.

There are several limitations that should be noted. First, although the overall patient cohort size was robust, stratification by pneumonia etiology resulted in smaller subgroups, which may limit statistical power for intergroup comparisons. Second, BAL sampling was performed based on clinical indications, potentially introducing bias due to variability in sampling times across individuals, although the overall distribution of samples across disease stages was generally balanced. Third, immune cells obtained from BAL may not fully capture the entire spectrum of alveolar or interstitial immune landscape. Fourth, while our CyTOF panel was comprehensive, several functional markers were excluded due to suboptimal antibody binding with available metal isotopes, possibly omitting relevant biological information. Finally, although cryopreservation enabled batched measurement and minimized technical variability, it may have affected cellular phenotypes compared to freshly processed samples.(94)

In summary, this work provides a comprehensive, protein-level atlas of alveolar immunity in SARF and reveals how dynamic, functionally specialized immune states, not gross cell proportions, encode patient trajectory. These data bridge the gap between pathogen-triggered injury and downstream immunopathology, offering mechanistic entry points for precision immunomodulatory therapies in pneumonia, ARDS, and broader causes of acute respiratory failure.

## Data Availability

All data produced in the present study are available upon reasonable request to the authors

## Declarations

## Ethics approval

The ethical boards of the participating hospitals approved the collection of data for the study purposes, ID AUMC 2020_065.

## Funding

This research was funded by an Amsterdam UMC fellowship in 2020 (no award/grant number) and the VIDI grant “Navigating treatment response in acute respiratory distress syndrome using a biological compass” (grant number: 09150172210004) to LDJB. Additional support was received from ZonMw under project number 10430102110011.

## Data sharing

De-identified participant data with data dictionary can be shared after approval of a proposal with a signed data access agreement and always in collaboration with the study group.

## Contribution statement

SZ, LSB performed the sample collection and processing, LSB and HBVD performed the clinical data collection, SJ and SZ performed the data analysis and wrote the first draft of the manuscript. LDJB, JW, TVDP contributed to the study concept and design. All authors provided feedback on earlier versions of the manuscript. All authors read and approved the final manuscript.

## Methods * STAR

### Key resources table

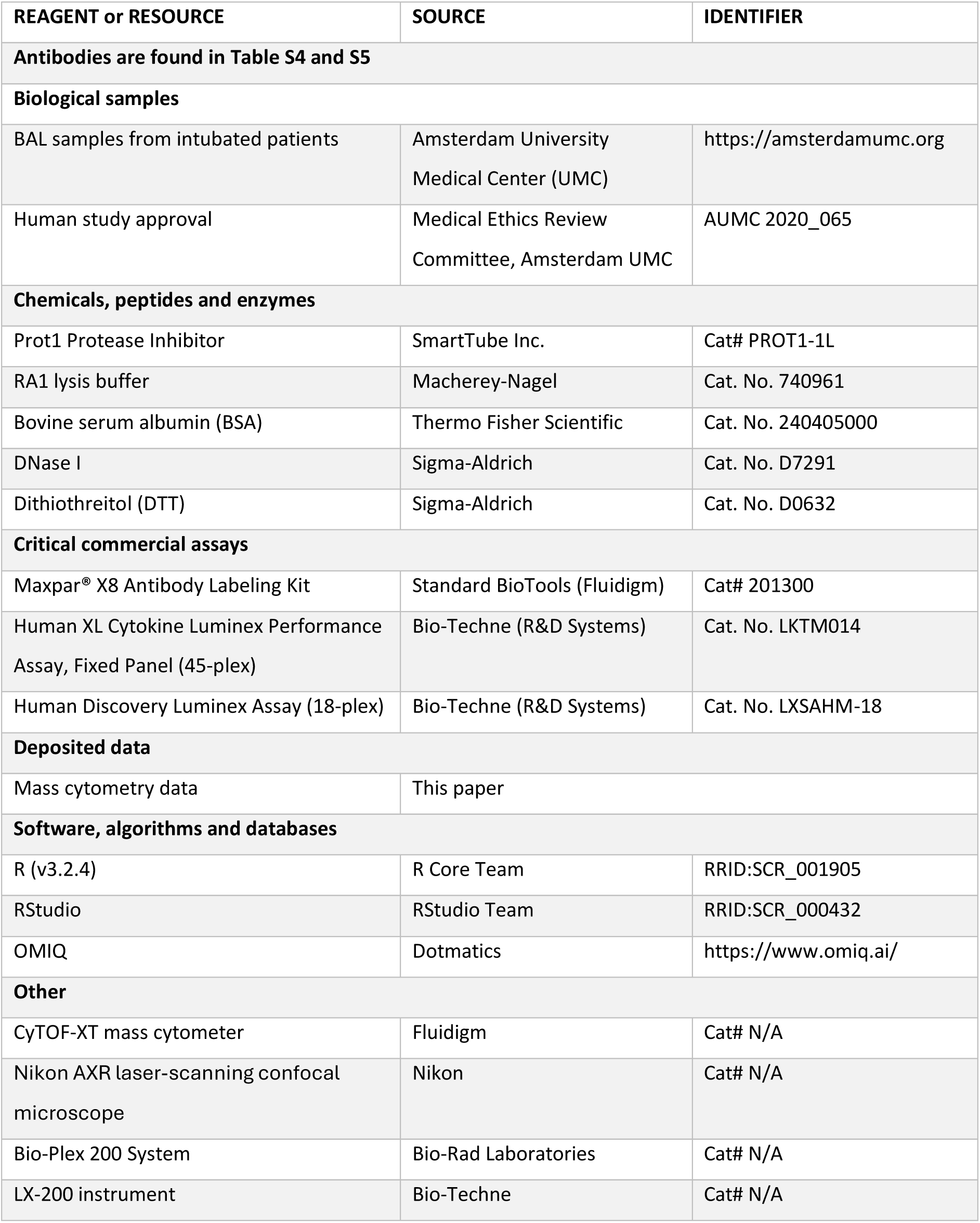

| REAGENT or RESOURCE | SOURCE | IDENTIFIER |
| --- | --- | --- |
| **Antibodies are found in Table S4 and S5** |  |  |
| **Biological samples** |  |  |
| BAL samples from intubated patients | Amsterdam University Medical Center (UMC) | https://amsterdamumc.org |
| Human study approval | Medical Ethics Review Committee, Amsterdam UMC | AUMC 2020_065 |
| **Chemicals, peptides and enzymes** |  |  |
| Prot1 Protease Inhibitor | SmartTube Inc. | Cat# PROT1-1L |
| RA1 lysis buffer | Macherey-Nagel | Cat. No. 740961 |
| Bovine serum albumin (BSA) | Thermo Fisher Scientific | Cat. No. 240405000 |
| DNase I | Sigma-Aldrich | Cat. No. D7291 |
| Dithiothreitol (DTT) | Sigma-Aldrich | Cat. No. D0632 |
| **Critical commercial assays** |  |  |
| Maxpar® X8 Antibody Labeling Kit | Standard BioTools (Fluidigm) | Cat# 201300 |
| Human XL Cytokine Luminex Performance Assay, Fixed Panel (45-plex) | Bio-Techne (R&D Systems) | Cat. No. LKTM014 |
| Human Discovery Luminex Assay (18-plex) | Bio-Techne (R&D Systems) | Cat. No. LXSAHM-18 |
| **Deposited data** |  |  |
| Mass cytometry data | This paper |  |
| **Software, algorithms and databases** |  |  |
| R (v3.2.4) | R Core Team | RRID:SCR_001905 |
| RStudio | RStudio Team | RRID:SCR_000432 |
| OMIQ | Dotmatics | https://www.omiq.ai/ |
| **Other** |  |  |
| CyTOF-XT mass cytometer | Fluidigm | Cat# N/A |
| Nikon AXR laser-scanning confocal microscope | Nikon | Cat# N/A |
| Bio-Plex 200 System | Bio-Rad Laboratories | Cat# N/A |
| LX-200 instrument | Bio-Techne | Cat# N/A |

### Study cohort and clinical data collection

This observational cohort study was conducted at Amsterdam University Medical Center, encompassing both AMC and VUmc locations in Amsterdam, the Netherlands. The study included all mechanically ventilated patients admitted to the ICU who underwent bronchoscopy with bronchoalveolar lavage (BAL) between August 20^th^ 2022 and May 8^th^ 2024. The diagnosis of pneumonia was based on a combination of clinical, radiological, and microbiological criteria. All clinical and microbiological findings were reviewed during multidisciplinary team discussions to ensure diagnostic accuracy and consistency with established guidelines. Patients with a high clinical likelihood of pneumonia were classified according to the identified pathogens, resulting in four categories: bacterial pneumonia (BP; common bacteria resulting in pneumonia), viral pneumonia (VP; Influenza A/B, RS virus, SARS-CoV2), bacterial + viral pneumonia (BVP; typically bacterial superinfection on a respiratory viral infection), other or culture-negative pneumonia (CNP, no apparent respiratory pathogen identified). Clinical data collection was conducted prospectively.

### Sample collection

As part of routine clinical care and following a standardized protocol, diagnostic bronchoscopy with BAL was performed weekly in patients showing no respiratory improvement. The decision to perform BAL was based on a consensus reached during multidisciplinary team meeting involving expert clinicians. During the bronchoscopy, 4×20ml 0.9% sodium chloride was inserted into single segments of the lungs and collected in fractions. The second fraction was used for our analysis, while the remaining fractions were used for microbiological diagnostics. To characterize immune profiles across different stages of disease progression, samples were stratified based on the timing of collection relative to intubation. Specifically, sample stages were defined as follows: early stage (0–4 days post-intubation), middle stage (5–10 days), and late stage (>10 days).

### Single cell isolation and preparation for mass cytometry

BAL samples were processed immediately upon collection. BAL sample was pooled into a clean 50 mL tube and centrifuged at 267 × g for 10 minutes at 8°C. The supernatant was transferred to a fresh tube without disturbing the cell pellet, which was then resuspended in 1 mL of PBK buffer, with additional volume added if excessive mucus was present. If the cell pellet was contaminated with mucus, it was treated with 5mM dithiothreitol (DTT, 1:1 vol/vol) and incubated on a shaker in a cold room for 15–30 minutes, with additional DTT added if needed until all mucus was dissolved. In cases of persistent cell clumping, DNase treatment (150 U/µL; starting at 1000:1 vol/vol) was applied and incubated for 10 minutes, repeated if necessary. If the pellet appeared red, red blood cell lysis was performed using 5 mL of 1× ELB, followed by incubation on ice for 10–15 minutes and centrifugation at 267 × g for 10 minutes at 8°C. The resulting cell pellet was gently resuspended in 1 mL of PBK buffer and kept on ice for cell counting. At least 500,000 viable cells from each sample were allocated into two 15 mL tubes for storage in PROT1 buffer according to the manufacturer’s instructions, ensuring stabilization of the proteome for subsequent mass cytometry analysis. All samples were stored at −80°C until further processing for CyTOF analysis. Additionally, cytospin slides were prepared per sample.

### Mass cytometry

#### Antibody preparation

All antibodies used in this study can be found in Table S1. Antibodies were purchased labeled or conjugated to their associated metals in-house. Antibody conjugation to heavy metal tags was done using the MaxPar Antibody Conjugation Kit (Fluidigm) according to the manufacturer’s protocol. Briefly, purified antibodies were subjected to buffer exchange into the kit-provided antibody labeling buffer using 50 kDa molecular weight cutoff filters. Following buffer exchange, the antibodies were partially reduced using TCEP (tris(2-carboxyethyl)phosphine) to expose free thiol groups. Concurrently, the metal-loaded polymer (MaxPar X8 polymer or Maxpar MCP9 Polymer based on the metal isotopes) was prepared by chelating a specific metal isotope. The reduced antibody and metal-loaded polymer were then incubated together to allow covalent binding through maleimide-thiol chemistry. Conjugated antibodies were subsequently purified to remove unconjugated polymers and other reagents using additional buffer exchanges. Final antibody concentrations were determined using a NanoDrop spectrophotometer, and conjugated antibodies were stored at 4°C in antibody stabilization buffer until use in mass cytometry staining panels.

#### Batch design and staining

BAL samples were distributed across 12 staining batches, balanced for age, mortality and infection using the *mixOmics* R package to minimize confounding. All antibodies were combined and aliquoted in order to maintain a uniform antibody mix for each batch. Each batch was stained separately, beginning with a surface antibody incubation in Cell Staining Buffer (Standard BioTools) for 30 minutes at room temperature. After surface staining, cells were permeabilized with methanol (−20°C, 20 minutes) and stained with intracellular antibodies for 30 minutes at room temperature.

#### Data acquisition

Following staining, cells were incubated with Cell-ID Intercalator-Ir (125 nM; Standard BioTools) and paraformaldahyde 1.6%(PFA) overnight. The following day, cells were washed in CAS+ (Standard BioTools) prior to acquisition on a CyTOF-XT mass cytometer (Standard BioTools) at an event rate of <400 events/second. Acquisition was performed sequentially across the 12 batches.

#### Data Preprocessing and Cleaning

Initial processing, including bead-based normalization, was performed using the embedded CyTOF-XT software suite. Normalized FCS files were exported and imported into the OMIC software package. Data were arcsinh-transformed with a cofactor of 5. Quality control was performed using PeacoQC to identify and exclude low-quality events.(95) Subsequent cleaning was carried out using Gaussian-based cleanup procedures.

Due to the debris-rich nature of the BAL samples, a two-step denoising process was applied. First, FlowSOM clustering was used to identify cellular populations.(20) Clusters displaying implausible or artifactual marker expression profiles were removed, while high-confidence clusters were retained. These were further refined based on DNA content using iridium intercalator intensity and event length metrics to select intact, live-like events.

#### Data analysis

Following identification of live single cells, a secondary round of FlowSOM clustering was performed to enable coarse stratification of broad immune cell lineages. This initial clustering distinguished major compartments: neutrophils, non-neutrophil myeloid cells (primarily macrophages and dendritic cells), and lymphoid cells. Clusters were manually annotated and cells were assigned to one of the three overarching categories based on canonical marker expression.

For downstream analysis, the dataset was split into two subsets. A total of 3 million neutrophil events were isolated into one subset. The second subset contained all of the non-neutrophil myeloid and lymphoid cells (2.5 million), which were jointly analyzed.

Each of the two subsets was clustered independently using PhenoGraph.(96) To aid in cluster annotation, marker enrichment modelling (MEM) was performed using the mem R package.(97) Dimensionality reduction was carried out using Uniform Manifold Approximation and Projection (UMAP) and t-distributed stochastic neighbor embedding (t-SNE). Cluster identities were defined by integrating MEM scores, low-dimensional visualization (UMAP and t-SNE), and manual gating. To facilitate metaclustering, MEM scores for each cluster were hierarchically clustered using the heatmapr function in R, applying Ward.D2 linkage. This hierarchical structure informed the consolidation of clusters into biologically and phenotypically meaningful metaclusters, representing distinct immune phenotypes. The specifics of cluster naming and corresponding annotations are shown in Supplemental materials.

#### Activation score calculation

To quantify activation states across major immune lineages, activation scores were computed at the single-cell level. For each cell, arcsinh-transformed expression values (preprocessed as described above) of markers associated with immune activation were selected as listed in Supplemental Table 2. Markers included in each pathway were determined through exploratory principal component analysis (PCA) and correlation plots of marker expression within each major cell type to identify markers that showed consistent co-expression patterns. Within each predefined cell lineage, activation marker intensities were first normalized, then summed and averaged by dividing by the number of markers included in the lineage-specific activation panel. These scores represent a composite activation state and were used for comparative analysis across conditions.

### Luminex assays

#### Cytokine assays

A comprehensive set of 70 biomarkers representative of the main pathophysiological pathways in ARDS (Table S1) was analyzed all at once in BAL supernatant. The biomarkers were measured by Luminex multiplex assay (R&D Systems, Abingdon, UK), using Bio-Plex 200 System (Bio-Rad Laboratories, Hercules, California, USA) and LX-200 instrument (Bio-Techne, Minneapolis, Minnesota, USA). The same assay kit was measured using the same equipment.

#### Data quality control

Data quality was assessed by evaluation of beads count (the number of replicates of a biomarker concentration measured). Measurements with a bead count lower than 25 were deemed a too low quality and were excluded. Values below the lowest point of the calibration curve were imputed with the lowest value (typically a concentration below biological relevance). Samples above the highest point of the calibration curve were extrapolated when possible (Table S7). Biomarkers with >25% of the values below the lower limit of quantification in BAL samples were considered unreliable and therefore excluded from the analyses.

### Thawing, immunolabeling, and flow-cytometric sorting

On the day of sorting, aliquots were thawed rapidly at 37 °C, diluted 1:10 in ice-cold PBS/2% FBS/2 mM EDTA, and centrifuged (400 g, 5 min). After two washes, 1 × 10⁶ cells were stained for 20 min at 4 °C with CD66b-FITC (clone G10F5, BioLegend), CD10-PE (clone HI10a, BD Biosciences) and CD16-APC (clone 3G8, BioLegend). Viable singlets were identified by forward- and side-scatter properties, doublet exclusion, and DAPI negativity. CD66b⁺ neutrophils were subdivided into CD10⁺CD16⁺, CD10⁻CD16⁺, CD10⁺CD16⁻ and CD10⁻CD16⁻ subsets and sorted on a FACSAria III (BD Biosciences) equipped with a 100 µm nozzle at 4 °C in purity mode. Post-sort analysis confirmed ≥95% purity for each fraction.

### Fluorescence microscopy

Sorted, PROT1-fixed cells (≈1 × 10⁵ per subset) were washed twice in PBS and incubated with DAPI (1 µg ml⁻¹, 5 min, room temperature). Ten- microliter drops of the suspension were placed on glass slides. Images were acquired on a Nikon AXR laser-scanning confocal microscope (Nikon Instruments) using a 40 × / 1.3 NA oil-immersion objective; laser power, detector gain, and offset were kept constant for all samples. Nuclear morphology (DAPI) and granule-membrane staining (CD66b-FITC) were assessed in at least 100 cells per subset.

### Research endpoints

We compared immune cell composition and activation across several key patient groups: 1) across patients with pneumonia and those without, with additional comparisons among pneumonia cases classified by causative pathogens; 2) across different disease stages: early, middle and late. To explore the relationship between alveolar immune responses and clinical outcomes, we assessed 3) associations between alveolar immune dynamics and mortality at 28-day after intubation. To investigate how alveolar cytokine levels reflect immune cell function, we assessed 4) the associations between immune cell profiles and alveolar cytokine levels; and 5) differences in cytokine levels between individual pneumonia subgroups and non-pneumonia cases.

### Statistical analysis

The non-parametric Wilcoxon rank sum test was utilized to compare the clinical characteristics, immune cell population frequencies, median protein expression values, immune activation scores between groups of interest. Kruskal-Wallis test was utilized to determine the significance among the groups of interest. The difference effect size of immune cell subset abundance and activation scores between individual pneumonia group and non-pneumonia group was assessed using Hedge’s *g*. The dynamic changes of alveolar immune cell subsets and activation scores were assessed using a negative binominal mixed effects model (*rstanarm* package) and logistical regression model (*lme4* package), respectively, in a longitudinal dataset. To evaluate the relationship between these immune dynamics and mortality, we applied a joint modeling approach (*rstanarm* package and *JM* package),(98, 99) which combined a longitudinal submodel and a Cox proportional hazards model. Spearman correlation analysis was conducted to assess the correlation in immune cell abundance and cytokine levels. All analyses were performed in R studio version 4.3.2 and the OMIQ software suite. Most of the plots were produced with the R package ggplot2.

## Online data supplement

### Tables

**Table S1.** Clinical characteristics of patients stratified by pneumonia etiology.

|  | Non-pneumonia patients | Pneumonia patients |  |  |  |
| --- | --- | --- | --- | --- | --- |
|  | Non-pneumonia<br>N=13 | Non-viral or<br>bacterial<br>pneumonia*<br>N=33 | Viral<br>pneumonia<br>N=14 | Bacterial<br>pneumonia†<br>N=21 | Bacterial +<br>viral<br>pneumonia<br>N=10 |
| **Patient characteristics** |  |  |  |  |  |
| Male, n (%) | 10 (76.9) | 20 (60.6) | 6 (42.9) | 18 (85.7) | 6 (60) |
| Age, median [IQR], years | 61 [55, 69] | 55 [45, 67] | 57 [39, 59] | 49 [29, 62] | 47 [35, 57] |
| BMI, median [IQR], kg/m<sup>2</sup> | 26.9 [23.1, 30.4] | 26.1 [21.3, 29.7] | 24.5 [23.1, 26.1] | 24.4 [22.2, 26.2] | 24.5 [20.2, 33.6] |
| Smoker, n(%) | 2 (16.7) | 5 (20) | 4 (36.4) | 7 (43.8) | 3 (37.5) |
| Days from hospital admission to ICU admission, median [IQR] | 1 [0, 4] | 3 [0, 14] | 0 [0, 3] | 0 [0, 1] | 0 [0, 1] |
| Days from ICU admission to intubation, median [IQR] | 0 [0, 3] | 0 [0, 1] | 0 [0, 1] | 0 [0, 0] | 0 [0, 0] |
| Admission type, n(%) |  |  |  |  |  |
| Surgical | 5 (38.5) | 6 (18.2) | 1 (7.1) | 9 (42.9) | 0 (0) |
| Medical | 8 | 28 | 13 | 12 | 10 |
| **Comorbidities, n(%)** |  |  |  |  |  |
| Immunocompromised conditions |  |  |  |  |  |
| Neutropenia | 0 (0) | 4 (12.5) | 1 (7.1) | 1 (4.8) | 1 (10) |
| Hematologic malignancy | 2 (14.3) | 6 (18.8) | 2 (15.4) | 1 (4.8) | 1 (10) |
| Allogeneic stem cell transplant | 1 (7.1) | 5 (15.6) | 1 (7.1) | 1 (4.8) | 0 (0) |
| Solid organ transplant | 1 (7.1) | 3 (9.4) | 0 (0) | 1 (4.8) | 1 (10) |
| Asthma | 0 (0) | 4 (12.5) | 3 (23.1) | 3 (15) | 0 (0) |
| COPD | 2 (14.3) | 1 (3.1) | 0 (0) | 1 (5) | 1 (10.0) |
| Interstitial lung disease | 3 (21.4) | 0 (0) | 1 (7.1) | 0 (0) | 0 (0) |
| **Cause of Acute Respiratory Failure\*** |  |  |  |  |  |
| ARDS | 8 (61.5) | 31 (93.9) | 14 (100) | 17 (81) | 9 (100) |
| Sepsis | 2 (15.4) | 13 (40.6) | 2 (15.4) | 7 (33.3) | 3 (30) |
| Atelectasis | 5 (38.5) | 16 (50.0) | 4 (30.8) | 11 (52.4) | 4 (40) |
| Aspiration | 1 (7.1) | 3 (9.4) | 2 (15.4) | 5 (23.8) | 1 (10) |
| Pulmonary embolism | 1 (7.1) | 2 (6.2) | 1 (7.7) | 2 (9.5) | 1 (10) |
| Cardiogenic pulmonary edema | 0 (0) | 4 (12.5) | 1 (7.7) | 4 (19) | 0 (0) |
| Emphysema | 1 (7.1) | 3 (9.4) | 2 (15.4) | 1 (4.8) | 0 (0) |
| COPD | 1 (7.1) | 0 (0) | 0 (0) | 0 (0) | 1 (10) |
| **Pneumonia pathogens, n(%)** |  |  |  |  |  |
| Virus |  |  |  |  |  |
| Influenza A | - | - | 8 (57.1) | - | 7 (70) |
| Influenza B | - | - | 1 (7.1) | - | 1 (10) |
| SARS-CoV-2 | - | - | 3 (21.4) | - | 1 (10) |
| RSV | - | - | 2 (14.3) | - | 1 (10) |
| Bacteria |  |  |  |  |  |
| Streptococcus | - | - | - | 0 (0) | 0 (0) |
| Klebsiella pneumoniae | - | - | - | 2 (9.5) | 0 (0) |
| Pseudomonas aeruginosa | - | - | - | 4 (19) | 1 (10) |
| Streptococcus pneumoniae | - | - | - | 0 (0) | 5 (50) |
| Staphylococcus aureus | - | - | - | 5 (23.8) | 4 (40) |
| Serratia | - | - | - | 4 (19) | 0 (0) |
| Morganella morganii | - | - | - | 4 (19) | 0 (0) |
| Other bacteria | - | - | - | 6 (28.6) | 1 (10) |
| **Characteristics at time of first BAL, median [IQR]** |  |  |  |  |  |
| Days from intubation to BAL sampling | 6 [3, 11] | 6 [1, 10] | 5 [3, 9] | 9 [5, 11] | 3 [1, 8] |
| Days from ICU admission to BAL sampling | 6 [1, 9] | 2 [1, 8] | 4 [1, 7] | 9 [6, 11] | 3 [1, 8] |
| SOFA score | 8 [7, 9] | 9 [7, 11] | 8 [7, 9] | 10 [7, 11] | 8 [7, 11] |
| Invasive MV | 2 (14.2) | 6 (18.8) | 1 (7.7) | 4 (19) | 1 (10) |
| PEEP, cmH<sub>2</sub>O | 8 [5, 12] | 8 [6, 10] | 10 [8, 12] | 10 [8, 12] | 8 [8, 10] |
| PaO<sub>2</sub>/FiO<sub>2</sub> | 125 [102, 206] | 155 [110, 204] | 114 [85, 179] | 129 [97, 187] | 142 [110, 179] |
| Dynamic compliance in mL/cm H<sub>2</sub>O | 28 [18, 49] | 22 [16, 29] | 25 [16, 74] | 28 [25, 35] | 25 [17, 54] |
| Ventilatory ratio | 1.8 [1.6, 2.6] | 1.7 [1.0, 2.2] | 1.8 [1.2, 2.1] | 1.7 [1.4, 2.1] | 1.3 [0.8, 2.3] |
| **Clinical outcomes** |  |  |  |  |  |
| ICU length of stay in days, median [IQR] | 17 [11, 24] | 24 [13, 35] | 17 [14, 23] | 23 [15, 32] | 39 [25, 50] |
| Days of mechanical ventilation, median [IQR] | 14 [8, 21] | 19 [8, 27] | 11 [7, 17] | 19 [13, 33] | 35 [17, 42] |
| ICU mortality, n (%) | 5 (38.5) | 18 (54.5) | 4 (28.6) | 3 (14.3) | 0 (0) |
| 28-day mortality, n (%) | 6 (46.2) | 11 (33.3) | 5 (35.7) | 1 (4.8) | 0 (0) |
| 90-day mortality, n (%) | 6 (46.2) | 18 (54.5) | 5 (35.7) | 3 (14.3) | 0 (0) |

\* Patients diagnosed with pneumonia but with negative pneumonia pathogens or unknown etiology.
† **Multiple bacterial species may teste positive in one patient at the same time.**
COPD = Chronic obstructive pulmonary disease; ARDS = Acute respiratory distress syndrome; BAL = Bronchoalveolar lavage; RSV = Respiratory syncytial virus; PCP = Pneumocystis pneumonia; VZV=Varicella zoster; CMV = Cytomegalovirus; HSV = Herpes simplex virus; EBV = Epstein-Barr virus; ICU = Intensive care unit.

**Table S2.**
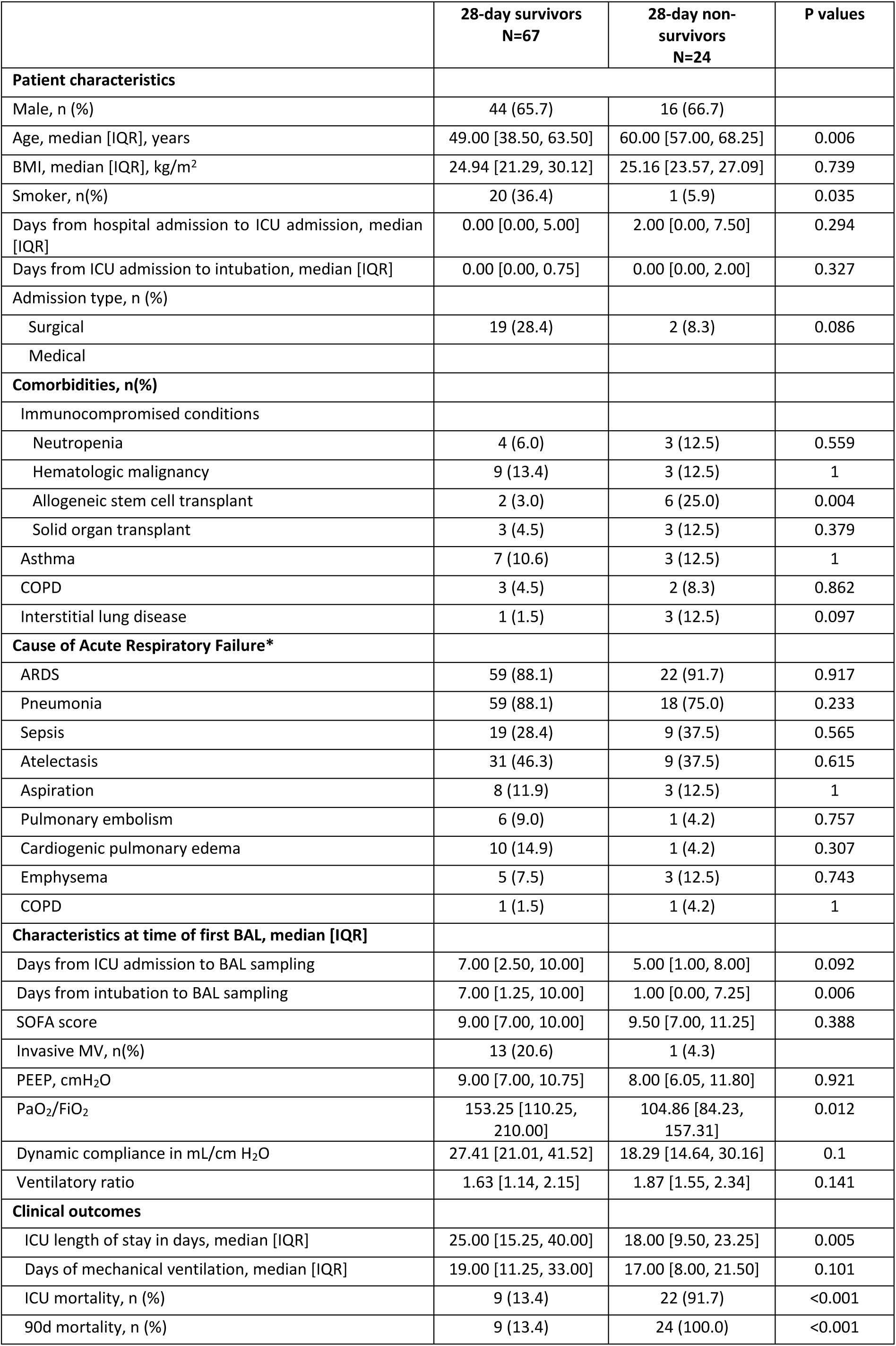
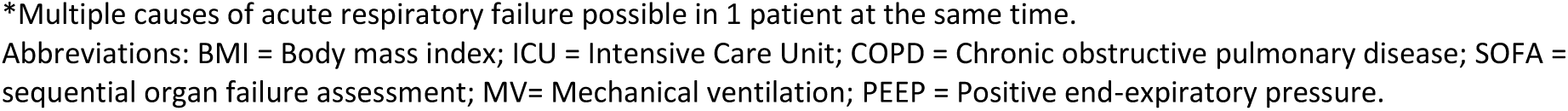
Clinical characteristics of patients stratified by 28-day survival.

|  | 28-day survivors<br>N=67 | 28-day non-survivors<br>N=24 | P values |
| --- | --- | --- | --- |
| **Patient characteristics** |  |  |  |
| Male, n (%) | 44 (65.7) | 16 (66.7) |  |
| Age, median [IQR], years | 49.00 [38.50, 63.50] | 60.00 [57.00, 68.25] | 0.006 |
| BMI, median [IQR], kg/m<sup>2</sup> | 24.94 [21.29, 30.12] | 25.16 [23.57, 27.09] | 0.739 |
| Smoker, n(%) | 20 (36.4) | 1 (5.9) | 0.035 |
| Days from hospital admission to ICU admission, median [IQR] | 0.00 [0.00, 5.00] | 2.00 [0.00, 7.50] | 0.294 |
| Days from ICU admission to intubation, median [IQR] | 0.00 [0.00, 0.75] | 0.00 [0.00, 2.00] | 0.327 |
| Admission type, n (%) |  |  |  |
| Surgical | 19 (28.4) | 2 (8.3) | 0.086 |
| Medical |  |  |  |
| **Comorbidities, n(%)** |  |  |  |
| Immunocompromised conditions |  |  |  |
| Neutropenia | 4 (6.0) | 3 (12.5) | 0.559 |
| Hematologic malignancy | 9 (13.4) | 3 (12.5) | 1 |
| Allogeneic stem cell transplant | 2 (3.0) | 6 (25.0) | 0.004 |
| Solid organ transplant | 3 (4.5) | 3 (12.5) | 0.379 |
| Asthma | 7 (10.6) | 3 (12.5) | 1 |
| COPD | 3 (4.5) | 2 (8.3) | 0.862 |
| Interstitial lung disease | 1 (1.5) | 3 (12.5) | 0.097 |
| **Cause of Acute Respiratory Failure\*** |  |  |  |
| ARDS | 59 (88.1) | 22 (91.7) | 0.917 |
| Pneumonia | 59 (88.1) | 18 (75.0) | 0.233 |
| Sepsis | 19 (28.4) | 9 (37.5) | 0.565 |
| Atelectasis | 31 (46.3) | 9 (37.5) | 0.615 |
| Aspiration | 8 (11.9) | 3 (12.5) | 1 |
| Pulmonary embolism | 6 (9.0) | 1 (4.2) | 0.757 |
| Cardiogenic pulmonary edema | 10 (14.9) | 1 (4.2) | 0.307 |
| Emphysema | 5 (7.5) | 3 (12.5) | 0.743 |
| COPD | 1 (1.5) | 1 (4.2) | 1 |
| **Characteristics at time of first BAL, median [IQR]** |  |  |  |
| Days from ICU admission to BAL sampling | 7.00 [2.50, 10.00] | 5.00 [1.00, 8.00] | 0.092 |
| Days from intubation to BAL sampling | 7.00 [1.25, 10.00] | 1.00 [0.00, 7.25] | 0.006 |
| SOFA score | 9.00 [7.00, 10.00] | 9.50 [7.00, 11.25] | 0.388 |
| Invasive MV, n(%) | 13 (20.6) | 1 (4.3) |  |
| PEEP, cmH<sub>2</sub>O | 9.00 [7.00, 10.75] | 8.00 [6.05, 11.80] | 0.921 |
| PaO<sub>2</sub>/FiO<sub>2</sub> | 153.25 [110.25, 210.00] | 104.86 [84.23, 157.31] | 0.012 |
| Dynamic compliance in mL/cm H<sub>2</sub>O | 27.41 [21.01, 41.52] | 18.29 [14.64, 30.16] | 0.1 |
| Ventilatory ratio | 1.63 [1.14, 2.15] | 1.87 [1.55, 2.34] | 0.141 |
| **Clinical outcomes** |  |  |  |
| ICU length of stay in days, median [IQR] | 25.00 [15.25, 40.00] | 18.00 [9.50, 23.25] | 0.005 |
| Days of mechanical ventilation, median [IQR] | 19.00 [11.25, 33.00] | 17.00 [8.00, 21.50] | 0.101 |
| ICU mortality, n (%) | 9 (13.4) | 22 (91.7) | <0.001 |
| 90d mortality, n (%) | 9 (13.4) | 24 (100.0) | <0.001 |

\*Multiple causes of acute respiratory failure possible in 1 patient at the same time.
Abbreviations: BMI = Body mass index; ICU = Intensive Care Unit; COPD = Chronic obstructive pulmonary disease; SOFA = sequential organ failure assessment; MV= Mechanical ventilation; PEEP = Positive end-expiratory pressure.

**Table S3.** Clinical characteristics of BAL sample collected at different stages of disease.

|  | Early stage<br>N=42 | Middle stage<br>N=44 | Late stage<br>N=40 |
| --- | --- | --- | --- |
| Days from intubation to BAL sampling, median [IQR], days | 1 [1, 4] | 8 [7, 10] | 17 [15, 24] |
| Days from ICU admission to BAL sampling, median [IQR], days | 1 [0, 2] | 8 [7, 9] | 17 [15, 23] |
| SOFA score | 9 [8, 11] | 9 [6, 11] | 9 [7, 11] |
| Type of ventilation support, n (%) |  |  |  |
| Volume control | 2 (4.8) | 0 (0) | 0 (0) |
| Pressure control | 21 (50) | 10 (22.7) | 14 (35) |
| Pressure support | 6 (14.3) | 26 (59.1) | 19 (47.5) |
| INTELLiVENT-ASV | 8 (19) | 5 (11.4) | 1 (2.5) |
| Pressure control and pressure support | 4 (9.5) | 1 (2.3) | 2 (5) |
| Ventilator parameters |  |  |  |
| PEEP, cmH2O | 9 [7, 12] | 9 [8, 10] | 8 [5, 10] |
| PaO2/FiO2 | 143.1 [97.2, 181.9] | 135.5 [104.3, 201.5] | 135.7 [118.7, 177.3] |
| Dynamic compliance in mL/cm H2O | 22 [15, 29] | 27 [16, 42] | 24 [19, 30] |
| Ventilatory ratio | 1.6 [1, 2.1] | 1.8 [1.2, 2.2] | 2 [1, 2.6] |

Abbreviations: BAL = bronchoalveolar lavage; ICU = Intensive Care Unit; SOFA = sequential organ failure assessment; PEEP = Positive end-expiratory pressure.

**Table S4.**
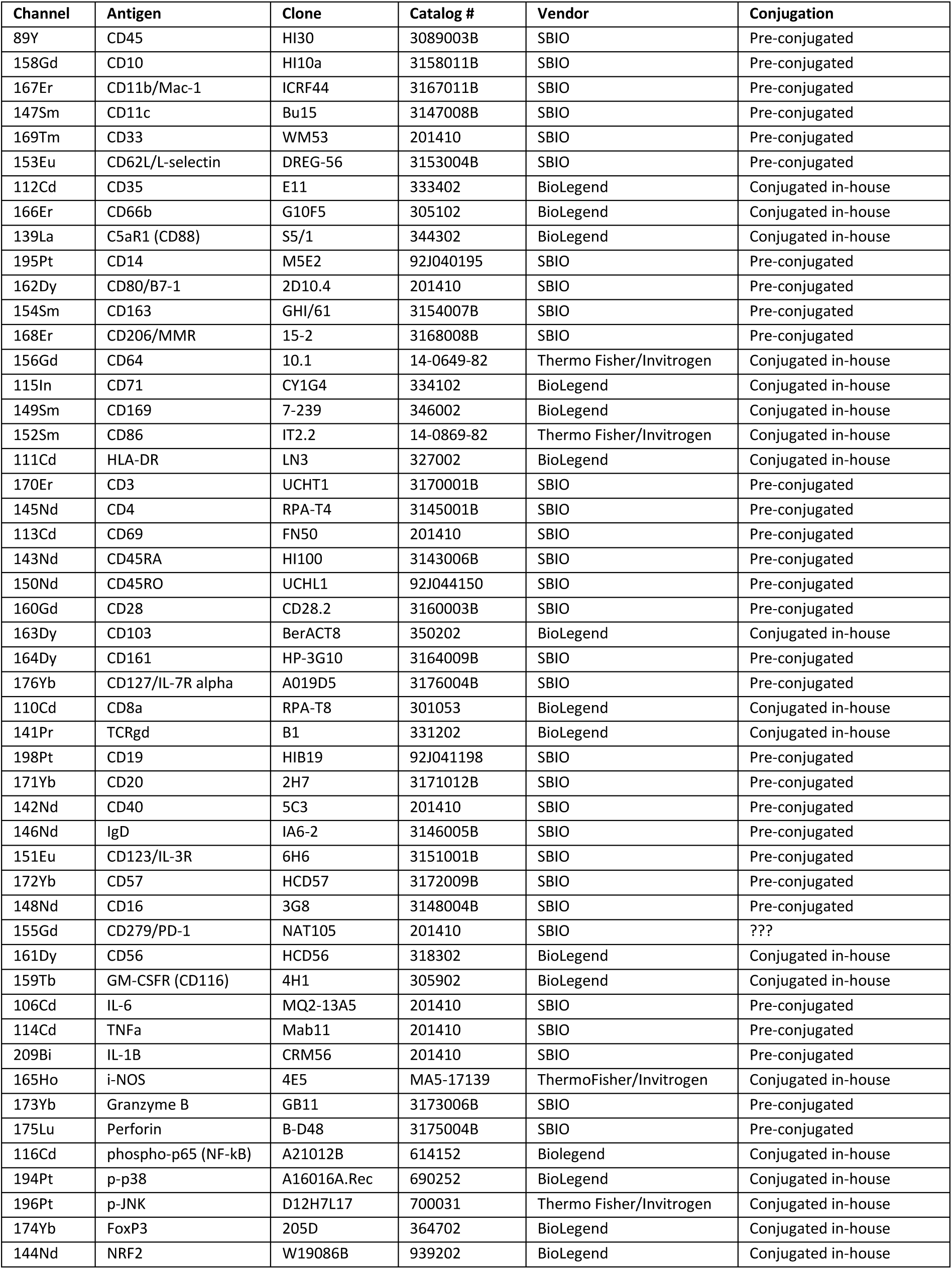
CYTOF antibody panel.

| Channel | Antigen | Clone | Catalog # | Vendor | Conjugation |
| --- | --- | --- | --- | --- | --- |
| 89Y | CD45 | HI30 | 3089003B | SBIO | Pre-conjugated |
| 158Gd | CD10 | HI10a | 3158011B | SBIO | Pre-conjugated |
| 167Er | CD11b/Mac-1 | ICRF44 | 3167011B | SBIO | Pre-conjugated |
| 147Sm | CD11c | Bu15 | 3147008B | SBIO | Pre-conjugated |
| 169Tm | CD33 | WM53 | 201410 | SBIO | Pre-conjugated |
| 153Eu | CD62L/L-selectin | DREG-56 | 3153004B | SBIO | Pre-conjugated |
| 112Cd | CD35 | E11 | 333402 | BioLegend | Conjugated in-house |
| 166Er | CD66b | G10F5 | 305102 | BioLegend | Conjugated in-house |
| 139La | C5aR1 (CD88) | S5/1 | 344302 | BioLegend | Conjugated in-house |
| 195Pt | CD14 | M5E2 | 92J040195 | SBIO | Pre-conjugated |
| 162Dy | CD80/B7-1 | 2D10.4 | 201410 | SBIO | Pre-conjugated |
| 154Sm | CD163 | GHI/61 | 3154007B | SBIO | Pre-conjugated |
| 168Er | CD206/MMR | 15-2 | 3168008B | SBIO | Pre-conjugated |
| 156Gd | CD64 | 10.1 | 14-0649-82 | Thermo Fisher/Invitrogen | Conjugated in-house |
| 115In | CD71 | CY1G4 | 334102 | BioLegend | Conjugated in-house |
| 149Sm | CD169 | 7-239 | 346002 | BioLegend | Conjugated in-house |
| 152Sm | CD86 | IT2.2 | 14-0869-82 | Thermo Fisher/Invitrogen | Conjugated in-house |
| 111Cd | HLA-DR | LN3 | 327002 | BioLegend | Conjugated in-house |
| 170Er | CD3 | UCHT1 | 3170001B | SBIO | Pre-conjugated |
| 145Nd | CD4 | RPA-T4 | 3145001B | SBIO | Pre-conjugated |
| 113Cd | CD69 | FN50 | 201410 | SBIO | Pre-conjugated |
| 143Nd | CD45RA | HI100 | 3143006B | SBIO | Pre-conjugated |
| 150Nd | CD45RO | UCHL1 | 92J044150 | SBIO | Pre-conjugated |
| 160Gd | CD28 | CD28.2 | 3160003B | SBIO | Pre-conjugated |
| 163Dy | CD103 | BerACT8 | 350202 | BioLegend | Conjugated in-house |
| 164Dy | CD161 | HP-3G10 | 3164009B | SBIO | Pre-conjugated |
| 176Yb | CD127/IL-7R alpha | A019D5 | 3176004B | SBIO | Pre-conjugated |
| 110Cd | CD8a | RPA-T8 | 301053 | BioLegend | Conjugated in-house |
| 141Pr | TCRgd | B1 | 331202 | BioLegend | Conjugated in-house |
| 198Pt | CD19 | HIB19 | 92J041198 | SBIO | Pre-conjugated |
| 171Yb | CD20 | 2H7 | 3171012B | SBIO | Pre-conjugated |
| 142Nd | CD40 | 5C3 | 201410 | SBIO | Pre-conjugated |
| 146Nd | IgD | IA6-2 | 3146005B | SBIO | Pre-conjugated |
| 151Eu | CD123/IL-3R | 6H6 | 3151001B | SBIO | Pre-conjugated |
| 172Yb | CD57 | HCD57 | 3172009B | SBIO | Pre-conjugated |
| 148Nd | CD16 | 3G8 | 3148004B | SBIO | Pre-conjugated |
| 155Gd | CD279/PD-1 | NAT105 | 201410 | SBIO | ??? |
| 161Dy | CD56 | HCD56 | 318302 | BioLegend | Conjugated in-house |
| 159Tb | GM-CSFR (CD116) | 4H1 | 305902 | BioLegend | Conjugated in-house |
| 106Cd | IL-6 | MQ2-13A5 | 201410 | SBIO | Pre-conjugated |
| 114Cd | TNFA | Mab11 | 201410 | SBIO | Pre-conjugated |
| 209Bi | IL-1B | CRM56 | 201410 | SBIO | Pre-conjugated |
| 165Ho | i-NOS | 4E5 | MA5-17139 | ThermoFisher/Invitrogen | Conjugated in-house |
| 173Yb | Granzyme B | GB11 | 3173006B | SBIO | Pre-conjugated |
| 175Lu | Perforin | B-D48 | 3175004B | SBIO | Pre-conjugated |
| 116Cd | phospho-p65 (NF-kB) | A21012B | 614152 | Biolegend | Conjugated in-house |
| 194Pt | p-p38 | A16016A.Rec | 690252 | BioLegend | Conjugated in-house |
| 196Pt | p-JNK | D12H7L17 | 700031 | Thermo Fisher/Invitrogen | Conjugated in-house |
| 174Yb | FoxP3 | 205D | 364702 | BioLegend | Conjugated in-house |
| 144Nd | NRF2 | W19086B | 939202 | BioLegend | Conjugated in-house |

**Table S5.** Flow cytometry antibody panel.

| **Antibody** | **Clone** | **Fluorochrome** | **Vendor** |
| --- | --- | --- | --- |
| CD10 | HI10a | Brilliant Violet 421™ | Biolegend |
| CD16 | 3G8 | PE-Cy7 | Biolegend |
| CD35 | E11 | APC | Biolegend |
| CD66b | G10F5 | FITC | Biolegend |
| CD45 | HI30 | Brilliant Violet 605™ | Biolegend |

**Table S6.** Frequency of cell subsets within each major cell type.

|  |  | Cluster percentages in each cell type, median [IQR] | Cluster percentages in each cell type, mean [SD] |
| --- | --- | --- | --- |
| **Neutrophils** | CD16bright Immature Neutrophil | 13.6 [6.0, 33.9] | 21.1 [19.8] |
|  | Degranulated Immature Neutrophil | 13.8 [7.2, 25.2] | 19.4 [16.4] |
|  | Apoptotic/dysfunctional Neutrophil | 7.8 [3.4, 13.8] | 11.3 [12.3] |
|  | CD123bright Immature Neutrophil | 2.3 [0.7, 6.4] | 7.5 [14.3] |
|  | CD16dim Mature Neutrophil | 6.7 [3.1, 10.0] | 9.1 [9.2] |
|  | Canonical Mature Neutrophil | 12.2 [4.6, 27.9] | 19.5 [19.7] |
|  | Resting Neutrophil | 7.8 [3.4, 13.8] | 12.2 [12.9] |
| **Myeloid non-neutrophil cells** | Monocyte-derived Macrophage | 28.2 [14.6, 45.0] | 29.9 [19.4] |
|  | Early Monocyte-derived Macrophage | 17.2 [6.5, 37.4] | 24.5 [22.8] |
|  | Resident Alveolar Macrophage | 13.2 [5.0, 30.4] | 20.9 [20.8] |
|  | Monocyte derived like Macrophage CD169high | 2.8 [1.2, 9.7] | 9.2 [16.2] |
|  | Mixed Macrophage | 10.2 [5.5, 20.0] | 15.3 [15.3] |
|  | pDCs | 0.1 [0.0, 0.3] | 0.3 [0.5] |
| **Lymphocytes** | Naive CD8 | 4.1 [2.2, 9.1] | 6.9 [6.8] |
|  | Naive CD4 | 0.3 [0.1, 1.0] | 1.2 [2.6] |
|  | CD4EM | 28.1 [15.5, 39.7] | 29.7 [17.4] |
|  | CD8EM | 29.2 [18.0, 49.3] | 26.3 [17.8] |
|  | CD8 NKT | 0.6 [0.2 - 1.7] | 1.5 [3.2] |
|  | Tissue resident CD4EM | 0.8 [0.4 - 2.3] | 2.1 [4.1] |
|  | Tissue resident CD8EM | 0.3 [0.1 - 1.0] | 2.9 [9.9] |
|  | CD8 CD161bright | 4.4 [2.3 - 7.4] | 5.8 [5.3] |
|  | Double negative T cell | 5.3 [2.4 - 10.8] | 9.6 [12.2] |
|  | B cells | 2.3 [0.85 - 4.9] | 4.1 [5.1] |
|  | NK cells | 7.2 [3.5 - 12.3] | 10.0 [9.2] |

**Table S7.** Alveolar cytokines and abbreviation.

| **Luminex biomarkers** | **Abbreviation** | **Alias** | **Category** |
| --- | --- | --- | --- |
| Matrix Metalloproteinase-1 | MMP-1 |  | Epithelial injury |
| Matrix Metalloproteinase-8 | MMP-8 |  | Epithelial injury |
| Tissue Inhibitor of Metalloproteinases-1 | TIMP-1 |  | Epithelial injury |
| Fibroblast Activation Protein Alpha | FAP-α | | Epithelial injury |
| Transforming Growth Factor Alpha | TGF-α | | Epithelial injury |
| Surfactant Protein D | SP-D |  | Epithelial injury |
| Receptor for Advanced Glycation End Products | RAGE |  | Epithelial injury |
| Periostin | Periostin | OSF | Epithelial injury |
| Platelet-Derived Growth Factor-AA | PDGF-AA |  | Epithelial injury |
| Platelet-Derived Growth Factor-AB/BB | PDGF-AB/BB |  | Epithelial injury |
| Interleukin-13 | IL-13 |  | Epithelial injury |
| Basic Fibroblast Growth Factor | bFGF | FGF-2 | Epithelial injury |
| Epidermal Growth Factor | EGF |  | Epithelial injury |
| Angiopoietin-1 | Angiopoietin-1 |  | Endothelial injury |
| Angiopoietin-2 | Angiopoietin-2 |  | Endothelial injury |
| Vascular Cell Adhesion Molecule-1 | VCAM-1 |  | Endothelial injury |
| Platelet Selectin | P-Selectin |  | Endothelial injury |
| Endothelial Selectin | E-Selectin |  | Endothelial injury |
| Vascular Endothelial Growth Factor | VEGF |  | Endothelial injury |
| Interleukin 2 | IL-2 |  | Immune cell activity |
| Interleukin 3 | IL-3 |  | Immune cell activity |
| Interleukin 4 | IL-4 |  | Immune cell activity |
| Interleukin 5 | IL-5 |  | Immune cell activity |
| Interleukin 7 | IL-7 |  | Immune cell activity |
| Interleukin 9 | IL-9 |  | Immune cell activity |
| Interleukin-12p70 | IL-12p70 |  | Immune cell activity |
| Interleukin-15 | IL-15 |  | Immune cell activity |
| Interferon Beta | IFN-β | | Immune cell activity |
| Interferon Alpha-2 | IFN-α2 | | Immune cell activity |
| Fms-like Tyrosine Kinase 3 Ligand | Flt3 ligand |  | Immune cell activity |
| Eotaxin | CCL11 | Eotaxin | Immune cell activity |
| CD40 Ligand | CD40 ligand | CD154 | Immune cell activity |
| Chemokine C-C motif ligand 22 | CCL22 |  | Immune cell activity |
| Regulated on Activation, Normal T Cell Expressed and Secreted | CCL5 | RANTES | Immune cell activity |
| Programmed Death-Ligand 1 | PD-L1 | CD274 | Immune cell activity |
| Macrophage Inflammatory Protein-3 Beta | CCL19 | MIP-3β | Immune cell activity |
| Macrophage Inflammatory Protein-3 Alpha | CCL20 | MIP-3α | Immune cell activity |
| Interferon Gamma-Induced Protein 10 | CXCL10 | IP-10 | Immune cell activity |
| Interleukin-1 Receptor Antagonist | IL-1RA |  | Anti-inflammatory |
| Interleukin 10 | IL-10 |  | Anti-inflammatory |
| Chemokine C-C motif ligand 18 | CCL18 |  | Anti-inflammatory |
| Ectonucleotide Pyrophosphatase | ENPP-2 | Autotaxin | Anti-inflammatory |
| Interleukin 6 | IL-6 |  | Pro-inflammatory |
| Interleukin 8 | CXCL8 | IL-8 | Pro-inflammatory |
| Macrophage Inflammatory Protein-1 Beta | CCL4 | MIP-1β | Pro-inflammatory |
| Macrophage Inflammatory Protein-1 Alpha | CCL3 | MIP-1α | Pro-inflammatory |
| Monocyte Chemoattractant Protein-1 | CCL2 | MCP-1 | Pro-inflammatory |
| Complement Component 9 | Complement C9 |  | Pro-inflammatory |
| Complement Component 5a | Complement C5a |  | Pro-inflammatory |
| Triggering Receptor Expressed on Myeloid Cells 1 | TREM-1 |  | Pro-inflammatory |
| Tumor Necrosis Factor Beta | Lymphotoxin-alpha | TNF-β | Pro-inflammatory |
| Tumor Necrosis Factor Alpha | TNF |  | Pro-inflammatory |
| Procalcitonin | Procalcitonin |  | Pro-inflammatory |
| Osteopontin | Osteopontin | SPP1 | Pro-inflammatory |
| Interleukin 17E | IL-25 |  | Pro-inflammatory |
| Interleukin 17A | IL-17 |  | Pro-inflammatory |
| Interferon Gamma | IFN-γ | | Pro-inflammatory |
| Growth-Regulated Oncogene Beta | CXCL2 | GRO-β | Pro-inflammatory |
| Growth-Regulated Oncogene Alpha | CXCL1 | GRO-α | Pro-inflammatory |
| Granulocyte-Macrophage Colony-Stimulating Factor | GM-CSF |  | Pro-inflammatory |
| Granulocyte Colony-Stimulating Factor | G-CSF |  | Pro-inflammatory |
| Interleukin-1 beta | IL-1β | | Pro-inflammatory |
| Interleukin-1 alpha | IL-1α | | Pro-inflammatory |
| Interleukin 13 | IL-33 |  | Pro-inflammatory |
| Tumor necrosis factor related apoptosis-inducing ligand | TRAIL |  | Programmed cell death |
| Tumor Necrosis Factor Receptor I | TNF-RI | TNFRSF1A | Programmed cell death |
| Granzyme B | Granzyme B |  | Programmed cell death |
| Thrombomodulin | Thrombomodulin |  | Coagulation and Fibrinolysis |
| von Willebrand Factor A2 Domain | vWF-A2 |  | Coagulation and Fibrinolysis |
| Urokinase-Type Plasminogen Activator | uPA |  | Coagulation and Fibrinolysis |

### Figures

**Figure S1.**
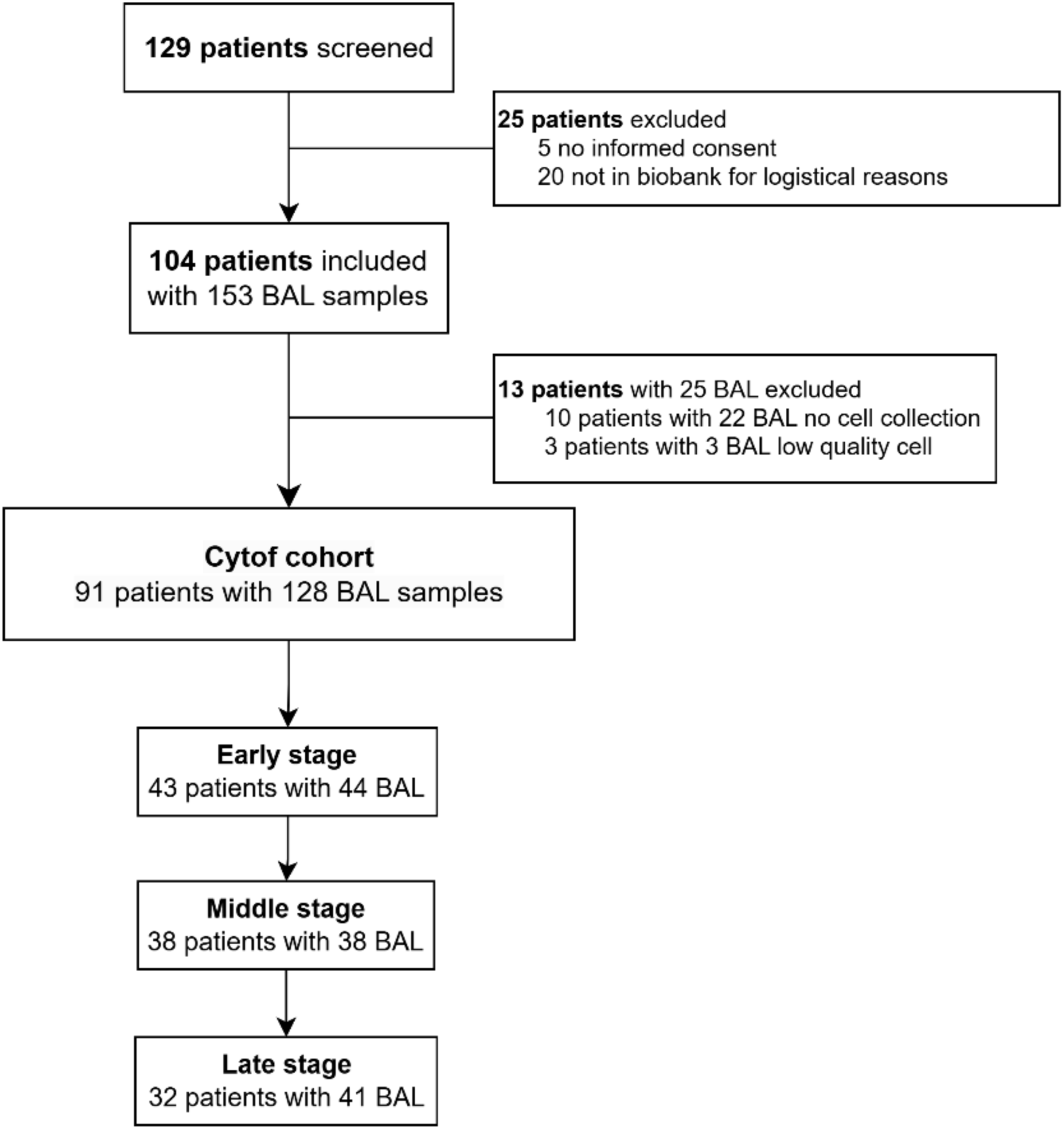
Flowchart of patient screen. BAL = bronchoalveolar lavage.

**Figure S2.**
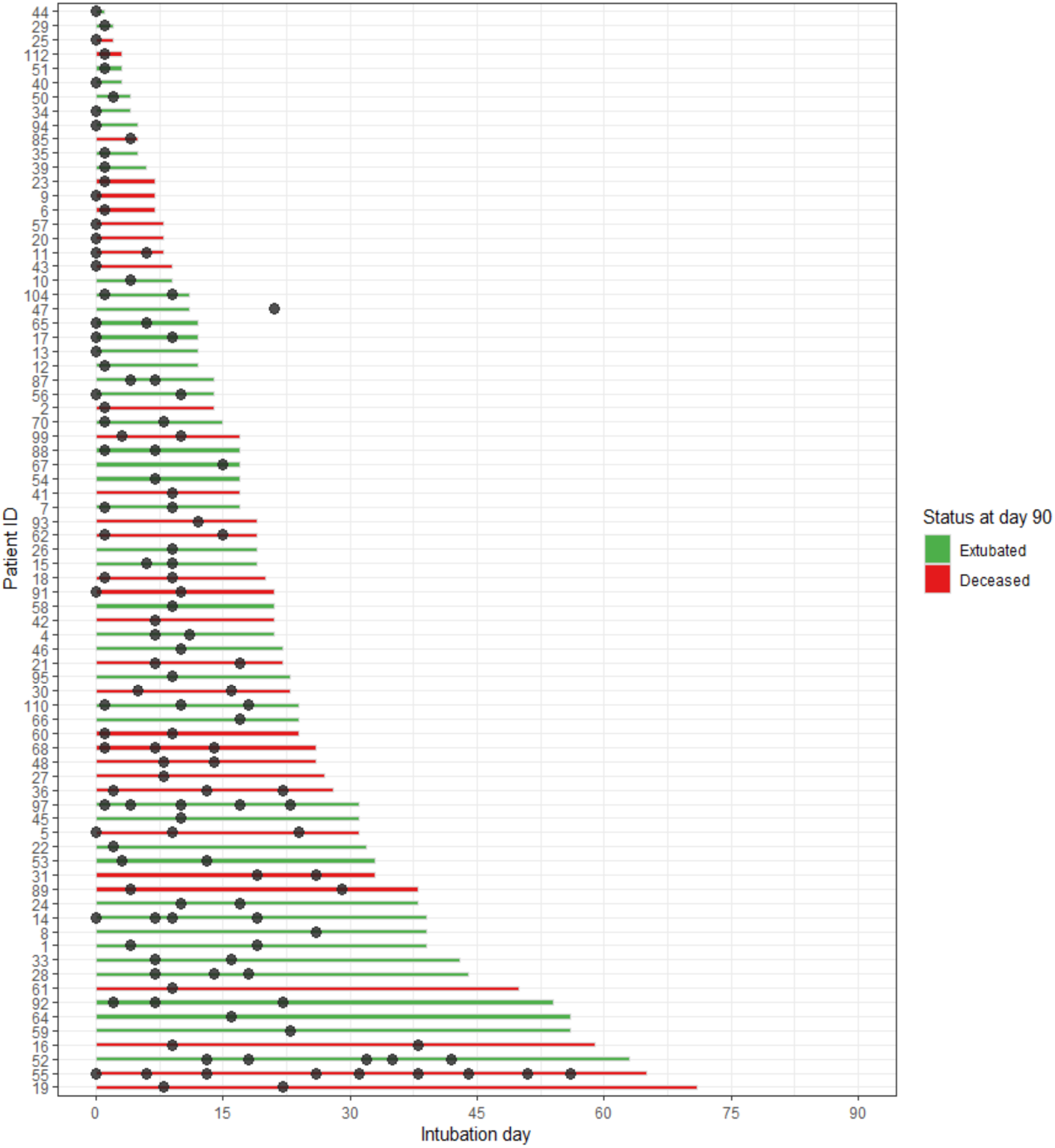
Swimmer plot of 91 patients included in this study. Time points of BAL samples are shown. Each line represents one subject in the study, from intubation until ICU discharge or death. The color of the line indicates the patients’ status at day 90 since intubation. Abbreviations*: BAL = bronchoalveolar lavage, ICU = intensive care unit, ID = identification*.

**Figure S3.**
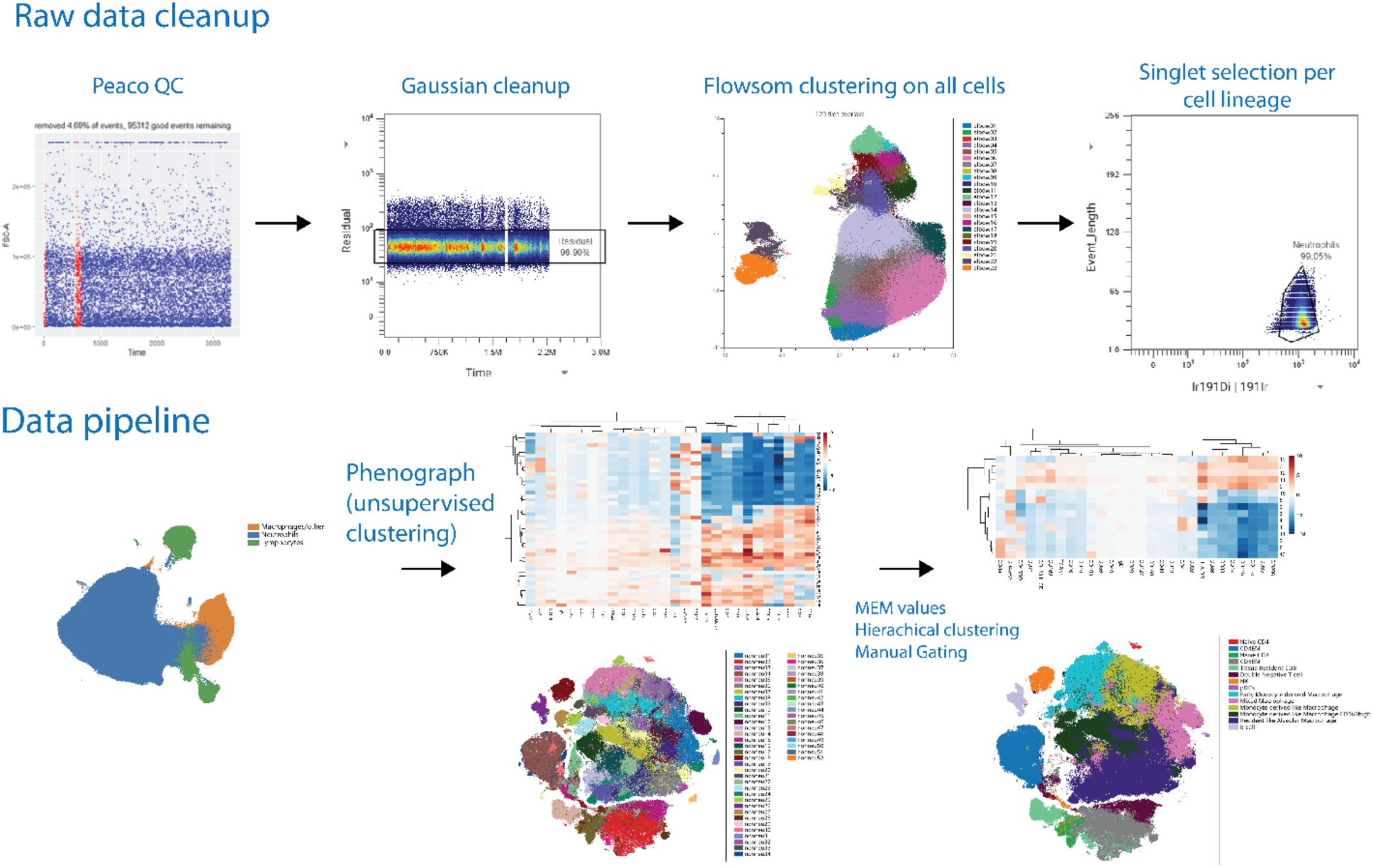
Overview of CyTOF data processing and analysis workflow. **Cleanup steps:** quality control (Peaco QC), Gaussian cleanup to remove acquisition artifacts, FlowSOM clustering on all cells, and singlet selection for lineage-specific analysis. **Data analysis pipeline:** visualization, Phenograph clustering, and generation of MEM scores followed by hierarchical clustering and manual gating to define immune cell subsets. Abbreviations*: QC=Quality control; MEM = marker enrichment modeling*.

**Figure S4.**
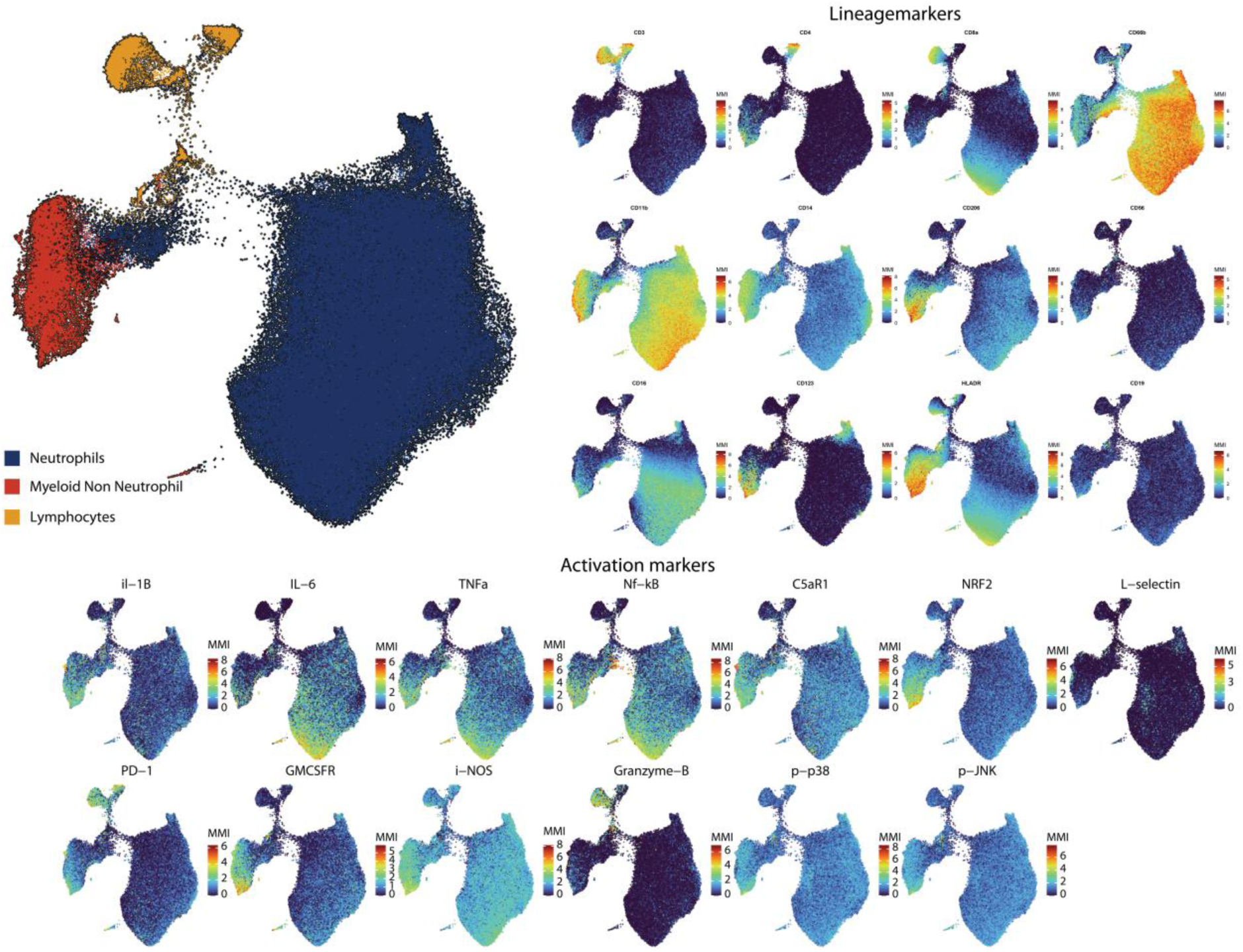
Identification of phenotypically distinct clusters in all immune cells. Pairwise Controlled Manifold Approximation Projection (PaCMAP) plot displaying clustering of alveolar immune cells, annotated into three main populations: neutrophils (blue), myeloid non-neutrophil cells (red), and lymphocytes (orange) based on lineage and functional marker expression.

**Figure S5.**
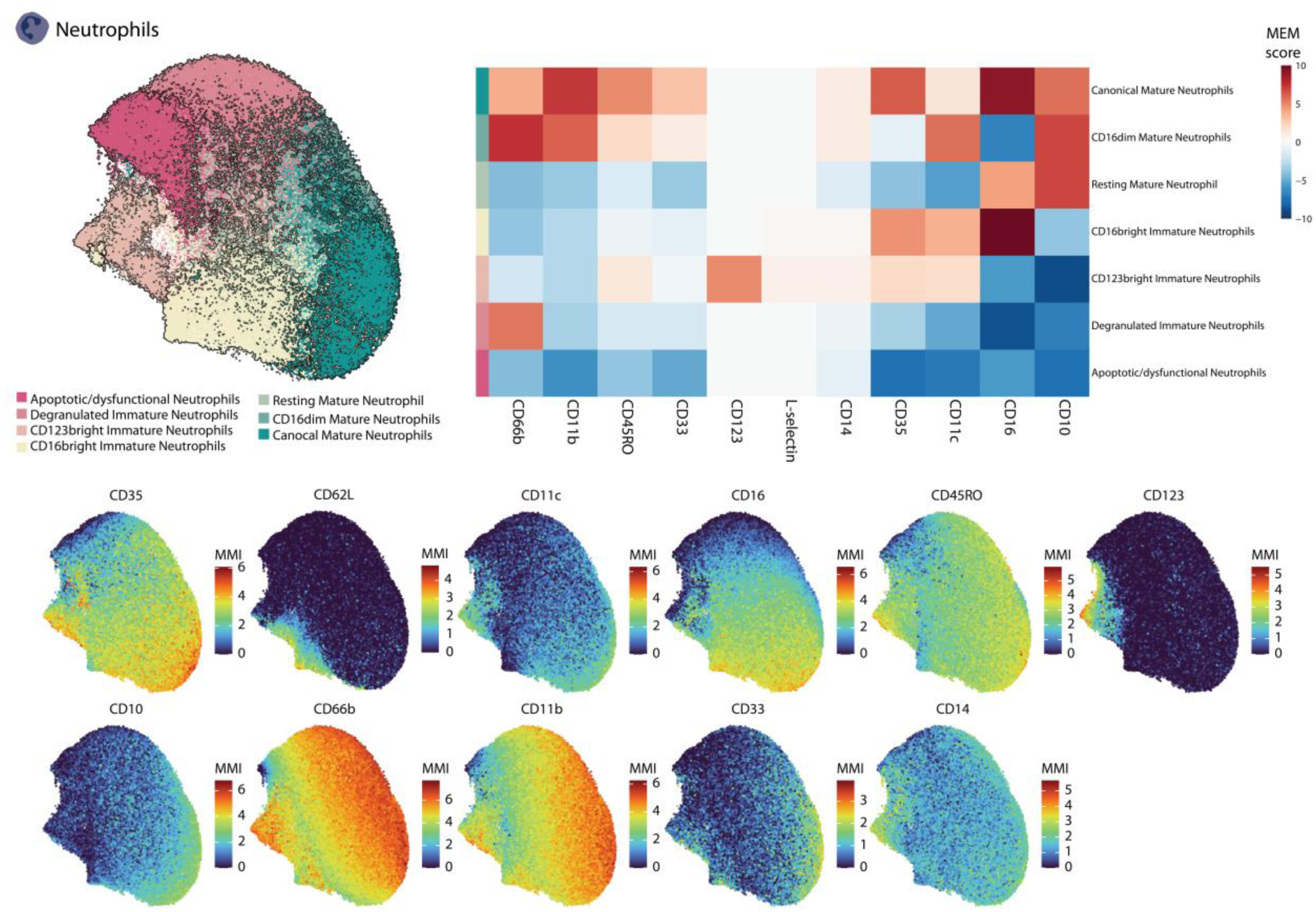
Identification of phenotypically distinct clusters in neutrophils. **(Top left)** Pairwise Controlled Manifold Approximation Projection (PaCMAP) plot showing clustering of neutrophils based on surface marker expression. **(Top right)** Heatmap of marker enrichment modeling (MEM) scores across neutrophil subsets highlights differential marker expression patterns distinguishing immature and mature phenotypes. **(Bottom)** PaCMAP plots overlaying median marker intensity (MMI) for selected surface markers.

**Figure S6.**
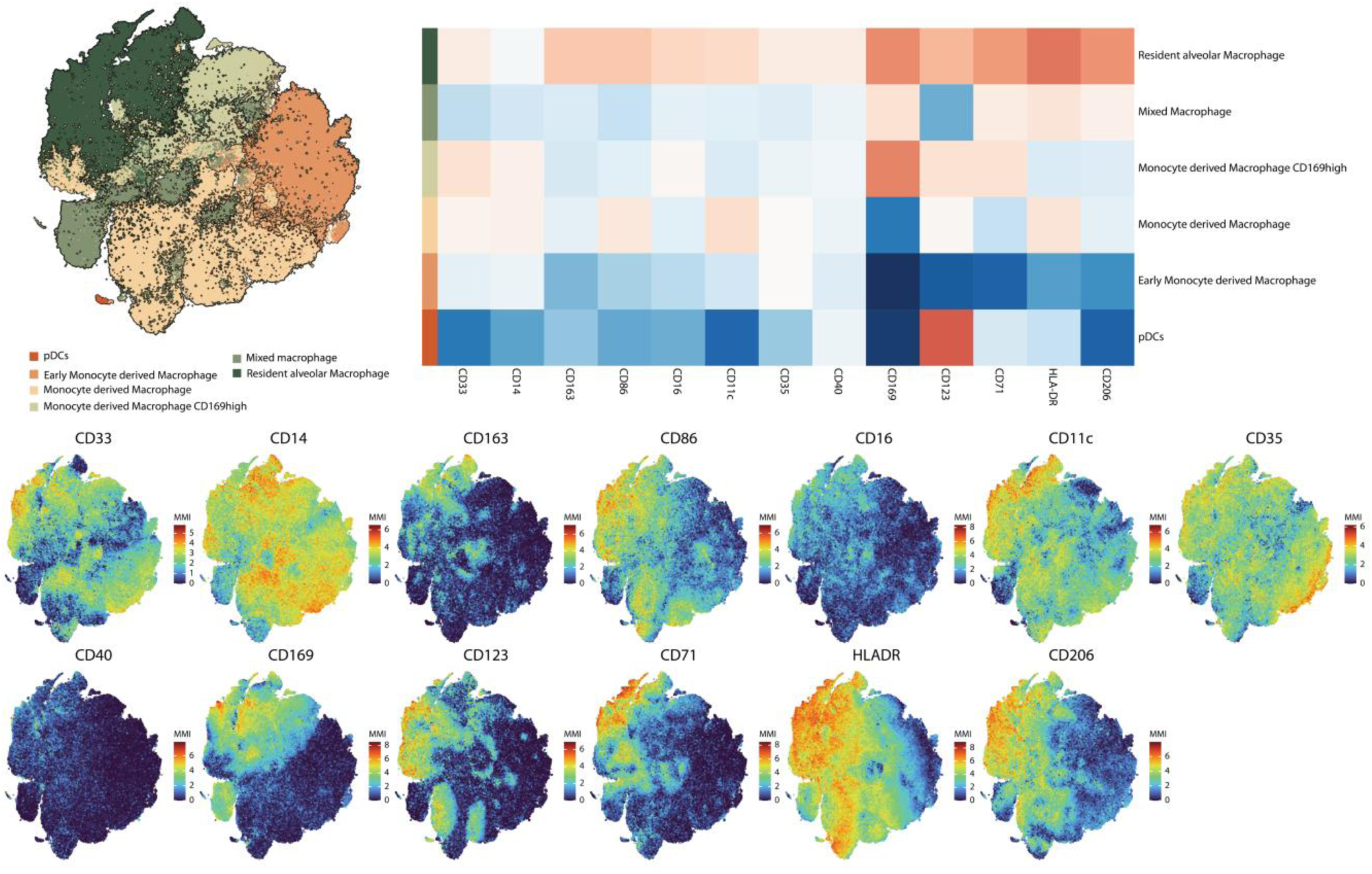
Identification of phenotypically distinct clusters in myeloid non-neutrophil cells. **(Top left)** T-distributed Stochastic Neighbor Embedding (t-SNE) plot showing clustering of myeloid non-neutrophil cells based on surface marker expression. **(Top right)** Heatmap of marker enrichment modeling (MEM) scores across myeloid non-neutrophil cell subsets highlights differential marker expression patterns distinguishing cell phenotypes. **(Bottom)** PaCMAP plots overlaying median marker intensity (MMI) for selected surface markers.

**Figure S7.**
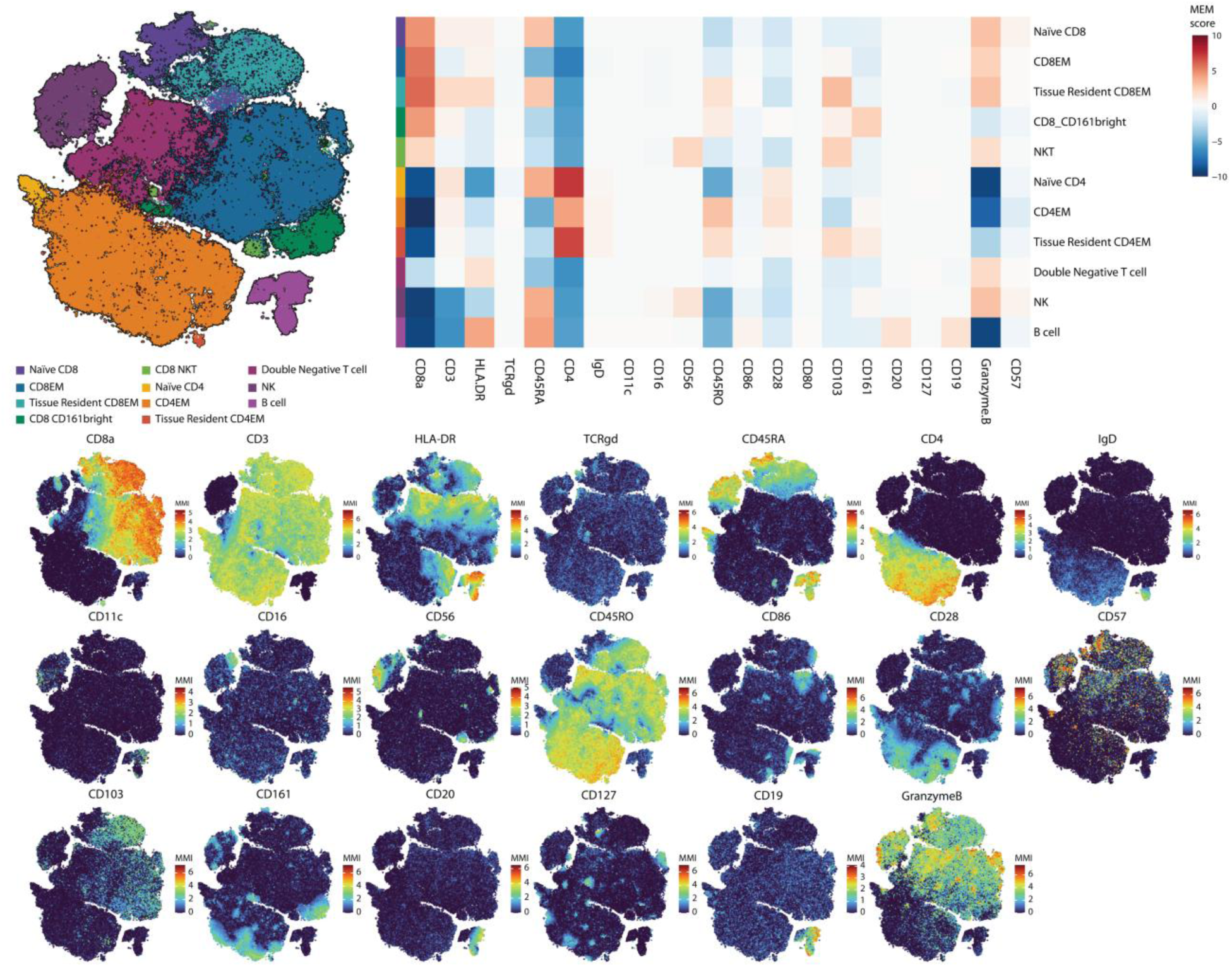
Identification of phenotypically distinct clusters in lymphocytes. **(Top left)** T-distributed Stochastic Neighbor Embedding (t-SNE) plot showing clustering of lymphocytes based on surface marker expression. **(Top right)** Heatmap of marker enrichment modeling (MEM) scores across lymphocyte subsets highlights differential marker expression patterns distinguishing cell phenotypes. **(Bottom)** PaCMAP plots overlaying median marker intensity (MMI) for selected surface markers.

**Figure S8.**
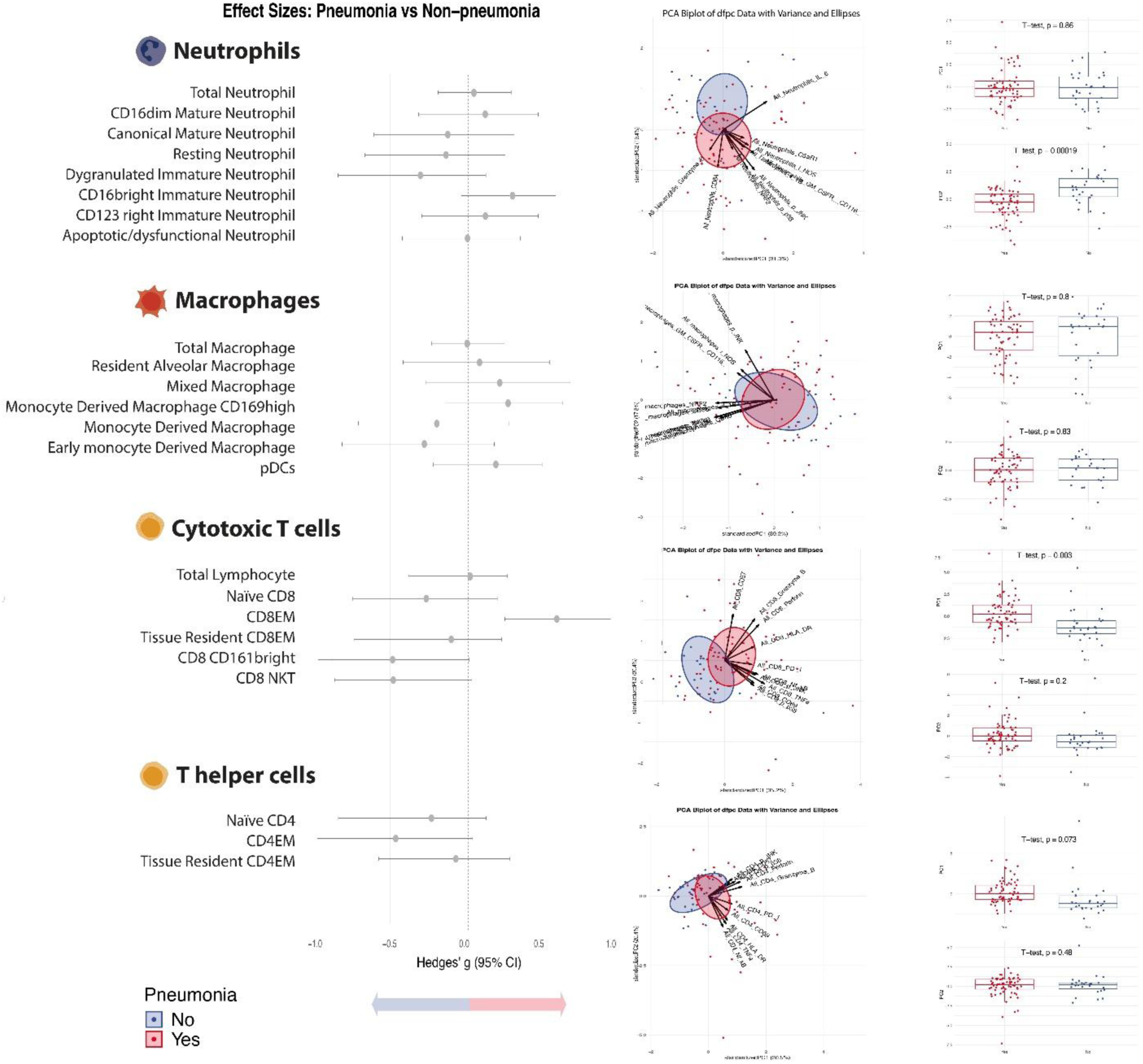
Alveolar immune cell subsets and distinct activation patterns between patients with and without pneumonia. **(Left)** Forest plot showing differences in cell subset abundance between patients with and without pneumonia. Each line indicates the 95% confident interval of the effect size (*Hedges’g*), with values centered relative to the non-pneumonia group. Gray points indicate non-significant differences. **(Right)** Principal component analysis (PCA) of functional marker expression that differentiates patient groups. Vectors represent the contribution and directionality of individual markers to group separation. Color boxes represent patient subgroups: red for pneumonia patients, blue for non-pneumonia patients.

**Figure S9.**
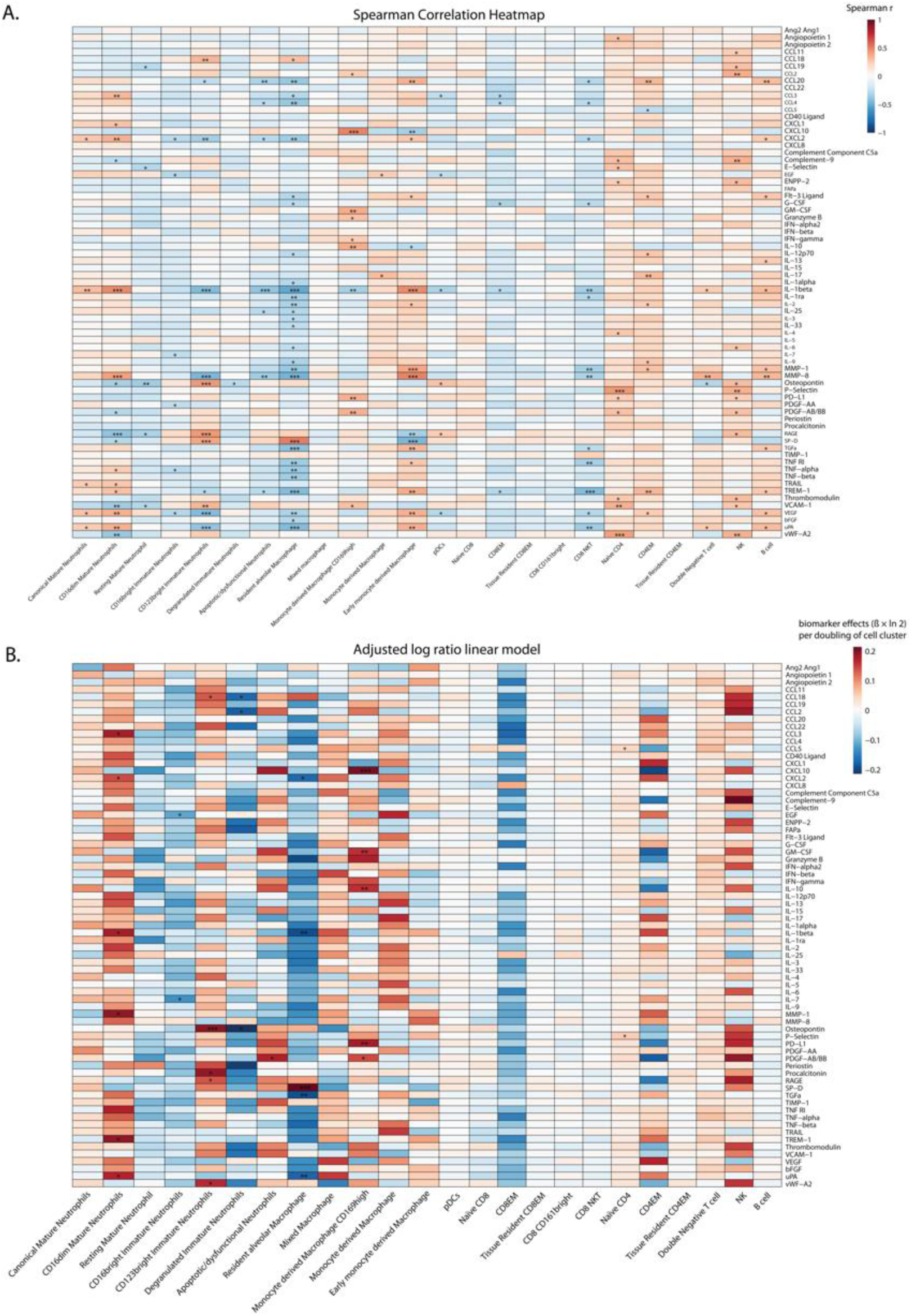
Associations between immune cell subsets and alveolar cytokines. **(A)** Spearman correlation heatmap depicting pairwise associations between immune cell subsets and alveolar cytokine levels. Colors indicate the strength and direction of the correlation coefficients. **(B)** Heatmap of adjusted log-ratio effect estimates from linear regression models assessing associations between immune cell subset frequencies and cytokine levels. Colors represent the magnitude and direction of the adjusted effects.

**Figure S10.**
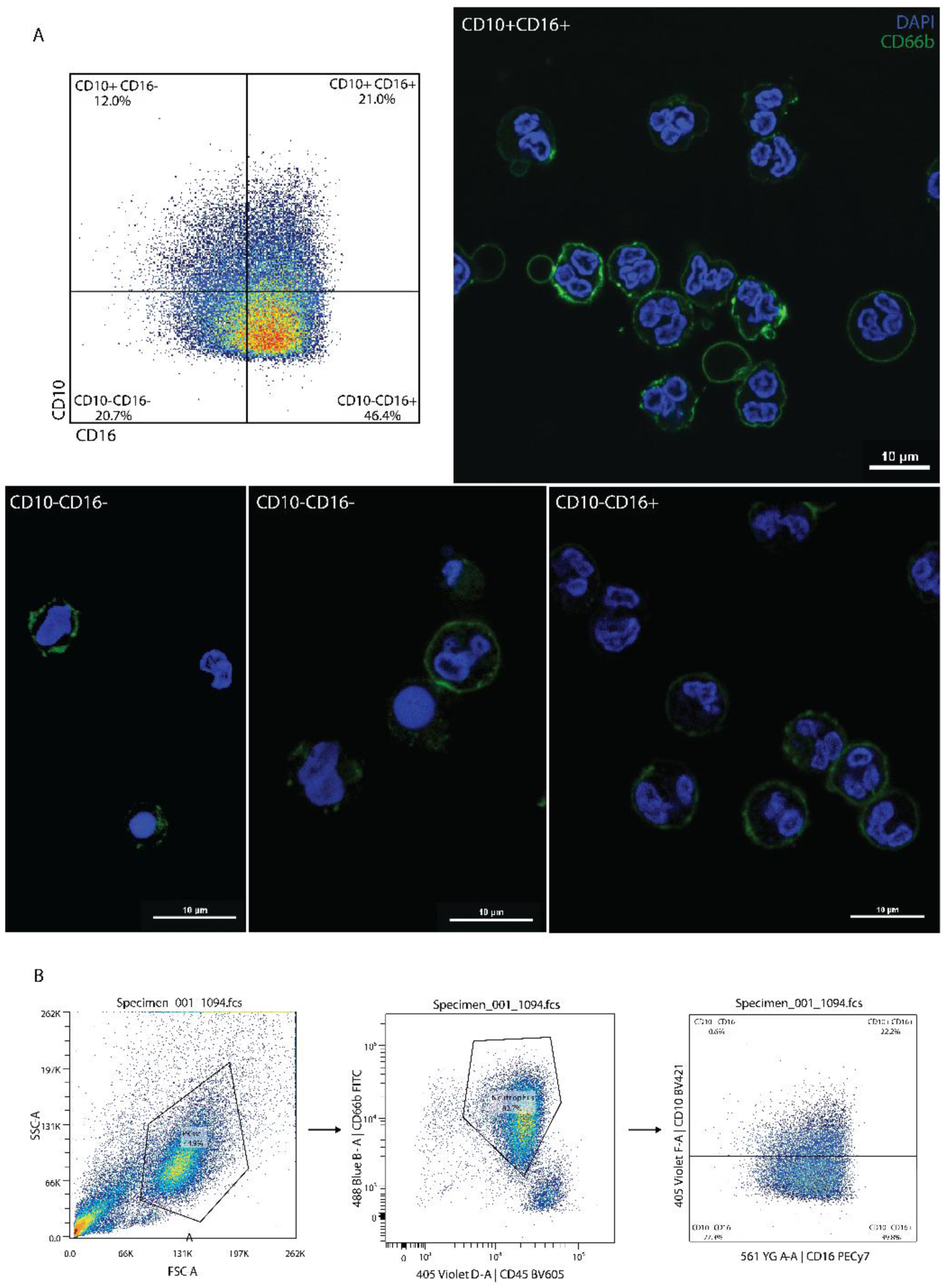
Phenotypic characterization and morphology of neutrophil subsets. **(A)** Flow cytometric analysis of neutrophil subsets based on CD10 and CD16 expression. **(B)** Gating strategy for neutrophil identification by flow cytometry.

**Figure S11.**
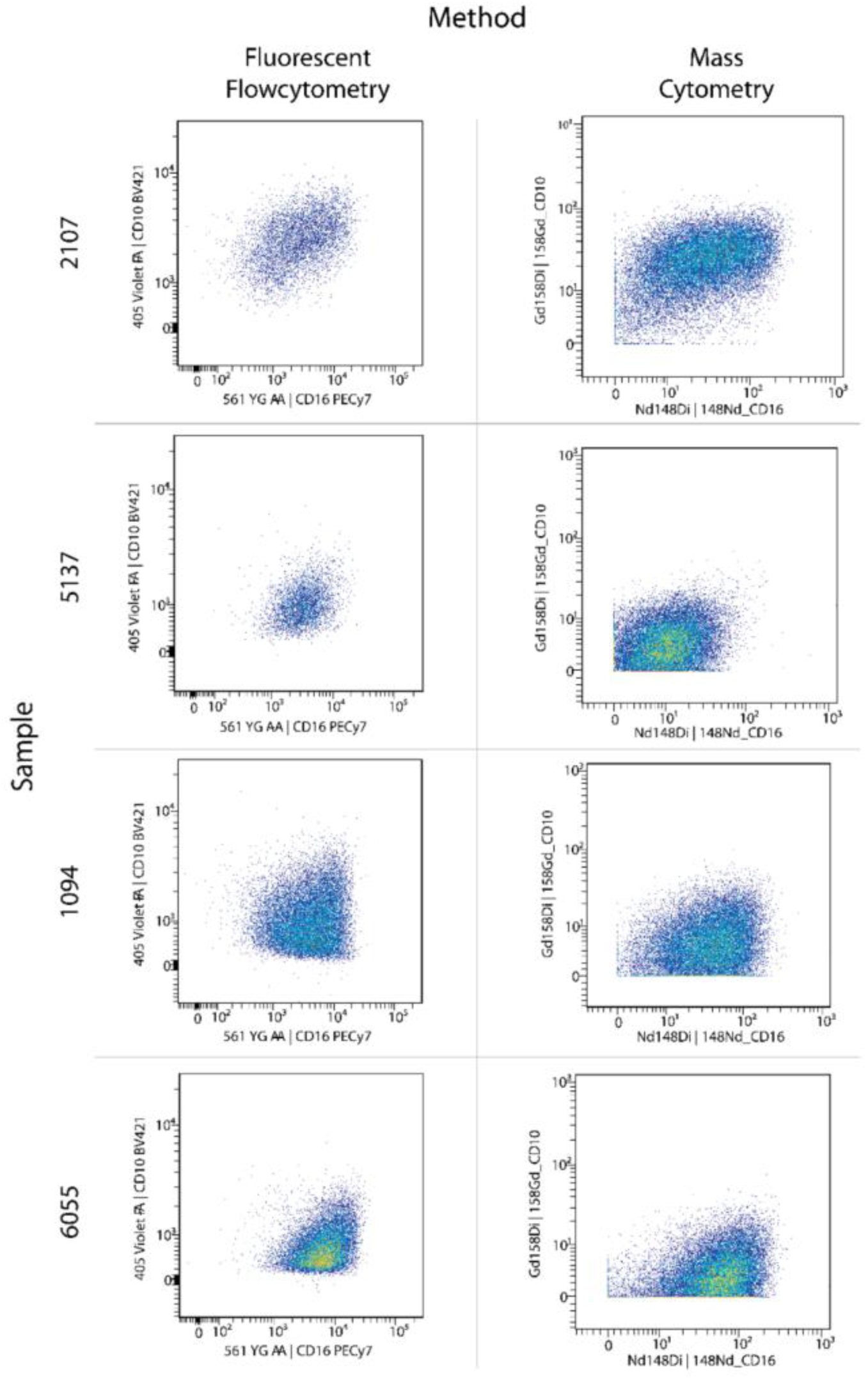
Comparison of CD10 and CD16 expression between flow cytometry and mass cytometry. Representative bivariate density plots showing CD10 versus CD16 expression in matched samples analyzed by fluorescent flow cytometry (left) and mass cytometry (CyTOF; right). Each row corresponds to an individual sample.

**Figure S12.**
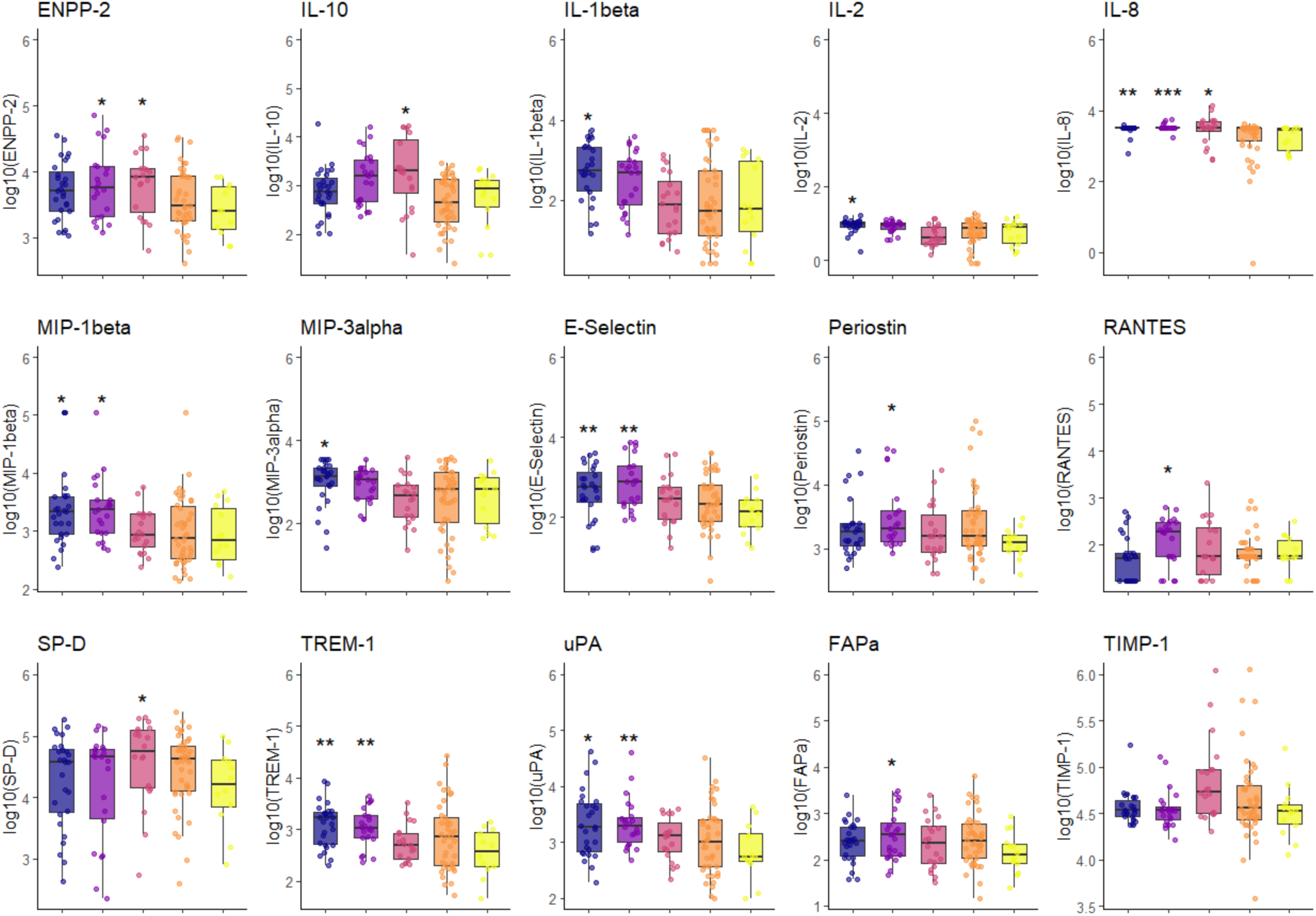
All significantly different cytokines among patient groups. Color boxes represent pneumonia subgroups: blue for bacterial pneumonia, purple for bacterial+viral pneumonia, pink for viral pneumonia, orange for culture-negative pneumonia, yellow for non-pneumonia. Abbreviations*: ENPP-2 = Ectonucleotide pyrophosphatase/phosphodiesterase 2; IL-10 = Interleukin 10; IL-1β = Interleukin 1beta; IL-2 = Interleukin 2; IL-8 = Interleukin 8; MIP-1β = Macrophage Inflammatory Protein 1 beta; MIP-3α = Macrophage Inflammatory Protein 3 alpha; E-Selectin = Endothelial-leukocyte adhesion molecule-1; RANTES = Regulated on Activation, Normal T cell Expressed and Secretedl; SP-D = Surfactant Protein D; TREM-1 = Triggering Receptor Expressed on Myeloid Cells 1; uPA = Urokinase-type Plasminogen Activator; FAPα = Fibroblast Activation Protein alpha; TIMP-1 = Tissue Inhibitor of Metalloproteinases*

## Notes

### Competing Interest Statement

The authors have declared no competing interest.

### Author Declarations

Biobank ethics committee of Academic Medical Center Amsterdam gave ethical approval for this work.

